# A Systematic Review and Meta-Analysis of the Impact of Tumour Mutation Burden on Survival Outcomes in Solid Tumours

**DOI:** 10.1101/2025.02.03.25321619

**Authors:** Aijia Meng, Alexander Yuile, Hao-Wen Sim, Subotheni Thavaneswaran, Humaira Noor, Jacky Yeung, Ashish Mehta, Joseph Powell, Ashraf Zaman

**Affiliations:** Garvan Institute of Medical Research, Sydney, Australia; Department of Medical Oncology, Royal North Shore Hospital, Sydney, Australia; Bill Walsh Translational Cancer Research Laboratory, The Kolling Institute, Sydney, Australia; School of Clinical Medicine, Faculty of Medicine and Health, University of New South Wales (UNSW), Sydney, Australia; The Kinghorn Cancer Centre, St Vincent’s Hospital, Sydney, Australia; NHMRC Clinical Trials Centre, University of Sydney, Sydney, Australia; Department of Medicine, Stanford Center for Biomedical Informatics Research (BMIR), Stanford University, Stanford, CA, USA; Department of Neurosurgery, Yale University School of Medicine, New Haven, CT, USA; UNSW Cellular Genomics Futures Institute, University of New South Wales (UNSW), Sydney, Australia

**Author notes:** **Corresponding author**: Dr Ashraf Zaman, Garvan Institute of Medical Research, Sydney, Australia.

**Keywords:** Meta-analysis, Tumour mutation burden (TMB), Overall survival (OS), Progression-free survival (PFS), Immunotherapy

## Abstract

**Background:** Tumour mutation burden (TMB) is an emerging pan-cancer biomarker with potential predictive value for immune checkpoint inhibitor (ICI) therapy outcomes. However, its prognostic significance remains inconsistent due to methodological variability and differing cut-off thresholds. This systematic review and meta-analysis evaluated the impact of TMB on overall survival (OS) and progression-free survival (PFS) across solid tumours.

**Methods:** Following PRISMA 2020 guidelines, we systematically searched PubMed, Scopus, ScienceDirect, and Cochrane databases for studies published between 2010 and 2024. Eligible studies reported hazard ratios (HRs) and 95% confidence intervals (CIs) comparing OS and PFS in high versus low TMB cohorts. Heterogeneity was assessed using the I² statistic, and publication bias via funnel plots and Egger’s test.

**Results:** A total of 5,278 patients across 28 studies were analysed. High TMB was significantly associated with improved OS and PFS, particularly in non-small cell lung cancer (OS: HR = 0.56), gastrointestinal cancers (OS: HR = 0.36), and advanced/recurrent tumours (OS: HR = 0.52). Survival benefits were most pronounced in ICI-treated patients, especially those receiving combination anti-PD-L1/PD-1 and anti-CTLA4 therapy (OS: HR = 0.47; PFS: HR = 0.50). Ultra-high TMB cases had superior outcomes (OS: HR = 0.44) compared to a universal 10 mut/Mb cut-off (OS: HR = 0.58). Variability in TMB measurement across sequencing platforms highlights the need for standardisation.

**Conclusion:** High TMB is a strong prognostic and predictive biomarker in ICI-treated cancers, yet methodological inconsistencies hinder clinical implementation. Standardising TMB assessment and refining clinically relevant thresholds are essential for optimising its role in precision oncology.

**PROSPERO registration number:** The protocol of this systematic review is registered on PROSPERO (CRD42024608809).

## Introduction

Immune checkpoint inhibitors (ICIs) have transformed cancer treatment, offering durable responses in several malignancies [1]. Over the past decade, ICIs have become a cornerstone of oncology research, driven by the success of programmed cell death-ligand 1 (PD-L1)/programmed cell death protein 1 (PD-1) inhibitors and cytotoxic T-lymphocyte antigen-4 (CTLA-4) inhibitors. These therapies reprogramme the tumour immune microenvironment by enhancing the effector functions of anti-tumour CD4+ and CD8+ T cells while attenuating regulatory T cell-mediated immunosuppression [2,3]. This dual mechanism impedes tumour evasion and enhances anti-tumour immune responses [4]. Despite their promise, response rates to ICIs vary widely across tumour types and even between patients with the same malignancy. Factors such as tumour heterogeneity, immune microenvironment differences, and intrinsic resistance mechanisms contribute to these discrepancies. Additionally, ICIs can cause severe immune-related toxicities, including colitis, hepatitis, and pneumonitis, necessitating reliable predictive biomarkers for patient selection.[5]

Tumour mutation burden (TMB) has emerged as a promising biomarker for ICI efficacy [6]. TMB quantifies the number of non-synonymous somatic mutations per megabase (mut/Mb) in tumour cells, measured using whole exome sequencing (WES) or targeted gene panels [7]. Theoretically, higher TMB reflects increased neoantigen generation, improving immune recognition and response to ICIs [6,7]. However, a consistent and comprehensive understanding of the relationship between TMB and survival outcomes remains elusive due to methodological variability and tumour-specific differences. This highlights the need for robust evidence synthesis to determine TMB’s prognostic value across solid tumours.

Inherited hypermutation results from genetic predispositions that impair DNA repair mechanisms. These predispositions produce highly immunogenic neoantigens that prime the immune system early in tumour development, often correlated with improved outcomes [8]. Conversely, acquired hypermutation, driven by environmental mutagens such as radiotherapy or chemotherapy, is less likely to yield favourable outcomes, particularly in advanced tumours [9,10]. These distinctions underscore the complexity of TMB as a biomarker.

Inflammatory characteristics also differ significantly across cancer types, with melanomas exhibiting a highly inflammatory microenvironment compared to gliomas’ predominantly anti-inflammatory milieu [11]. Such variability complicates cross-tumour analyses, leading many studies to focus on specific tumour types instead. While this site-specific approach yields valuable insights, it limits understanding of TMB’s broader predictive value across cancers. Previous meta-analyses have often focused on single tumour types or limited sample sizes, resulting in an incomplete understanding of TMB’s predictive capabilities across diverse malignancies.

This meta-analysis synthesised data from 28 studies encompassing 5,278 patients with solid tumours. By comparing overall survival (OS) and progression-free survival (PFS) between high and low TMB cohorts, the study investigated key sources of heterogeneity affecting TMB’s prognostic relevance. We analysed cancer type, treatment regimen, TMB threshold, and disease stage to provide insights into the mechanisms underlying TMB’s predictive capabilities. The findings aim to inform patient stratification strategies and refine therapeutic decision-making, advancing the clinical application of TMB as a biomarker.

## Method

### Literature search

This study focuses on the impact of TMB on survival outcomes across solid tumour studies. Research articles published from 1 January 2010 to 19 August 2024 were identified on PubMed, Scopus, ScienceDirect, and Cochrane databases. The starting year of 2010 was selected because TMB as a biomarker was not widely studied earlier, and the search end date of 19 August 2024 reflects the most recent studies available at the time of analysis. The search strategy used a combination of free-text terms and controlled vocabulary. Only articles published in English were included. Keywords used for the literature search included “TMB,” “tumour mutation burden,” “tumor mutation burden,” “tumour mutational burden,” “tumor mutational burden,” “survival,” “overall survival,” and “progression-free survival” (Supplementary File). Then, we manually reviewed the reference lists in these articles to identify any additional relevant publications.

### Inclusion and exclusion criteria

The inclusion criteria included: 1) Patients with a pathological diagnosis of a solid tumour. 2) The association between TMB value and at least one survival outcome indicator (overall survival [OS] or progression-free survival [PFS]) was reported. 3) Patients stratified by TMB cut-off into high and low TMB cohorts. 4) Hazard ratio (HR) values and 95% confidence intervals (CIs) for high versus low TMB cohorts were provided. 5) Studies published in English.

The exclusion criteria were: 1) Non-human studies, in vivo and in vitro experiments, case reports, conference abstracts, and duplicate publications. 2) Studies not measuring TMB from tumour tissue samples (e.g., studies using circulating tumour DNA or alternative biomarkers). 3) Studies with inaccessible full text. 4) Studies with unclear methodologies, insufficient TMB, or survival data for endpoints. 5) Meta-analyses with overlapping publications.

### Data extraction

Two researchers independently extracted data, with a third reviewer resolving conflicts through discussion and consensus. Extracted data included the study title, first author, year of publication, study country, data sources, TMB measurement method, cancer type, treatment regimen, disease stage, patient demographics, survival outcomes, HR for TMB-high versus TMB-low groups, and corresponding 95% CIs and *p*-values. We also recorded whether TMB cut-off values were predefined or varied across studies.

### Statistical analysis

All statistical analyses were performed using the “meta” and “metafor” packages in R (Version 4.4.1). HRs and 95% CIs were extracted to compare TMB-high versus TMB-low groups. Pooled HRs, summary effect size, 95% CI, and *p*-value were calculated using both fixed- and random-effects models, with results presented under random-effects models to account for expected heterogeneity. Heterogeneity was assessed using Cochran’s *Q*-test and quantified by the I-squared (I²) statistic. Thresholds for heterogeneity were defined as I² < 50% (low), 50%– 75% (moderate), and >75% (high), following established guidelines. To explore sources of heterogeneity, subgroup analyses were conducted by cancer type, treatment regimen, TMB cut-off, and disease stage. Publication bias was visualised using funnel plots and quantified using Egger’s test, where *p* > 0.05 indicates no evidence of publication bias.

## Results

### Study Selection and Characteristics

This study reviewed the survival outcomes of patients with solid tumours in relation to TMB, following PRISMA guidelines [12]. The initial search identified 2,511 potentially relevant articles, of which 28 studies published between January 2010 and August 2024 were included in the final analysis. These studies comprised a total of 5,278 cases across 11 cancer types, including 3,008 non-small cell lung cancers (NSCLC), 680 melanomas, 574 gastrointestinal (GI) cancers, 151 head and neck cancers, 136 penile squamous cell carcinomas (SCC), 99 gliomas, 54 ovarian cancers (OVC), 46 breast cancers, 45 cervical cancers, 31 osteosarcomas, and seven endometrial cancers. Treatment modalities included immunotherapy in 18 studies, chemotherapy in seven, and radiotherapy in one study. The study selection process is detailed in **Figure 1**, and the characteristics of included studies are summarised in **Table 1**. The details of the selected studies included in this meta-analysis, including study design, sample size, and key characteristics, are summarised in **Supplementary Table S1**, while the pooled patient data, including the distribution of cancer types and the number of patients in each category, are presented in **Supplementary Table S2**.

**Figure 1.**
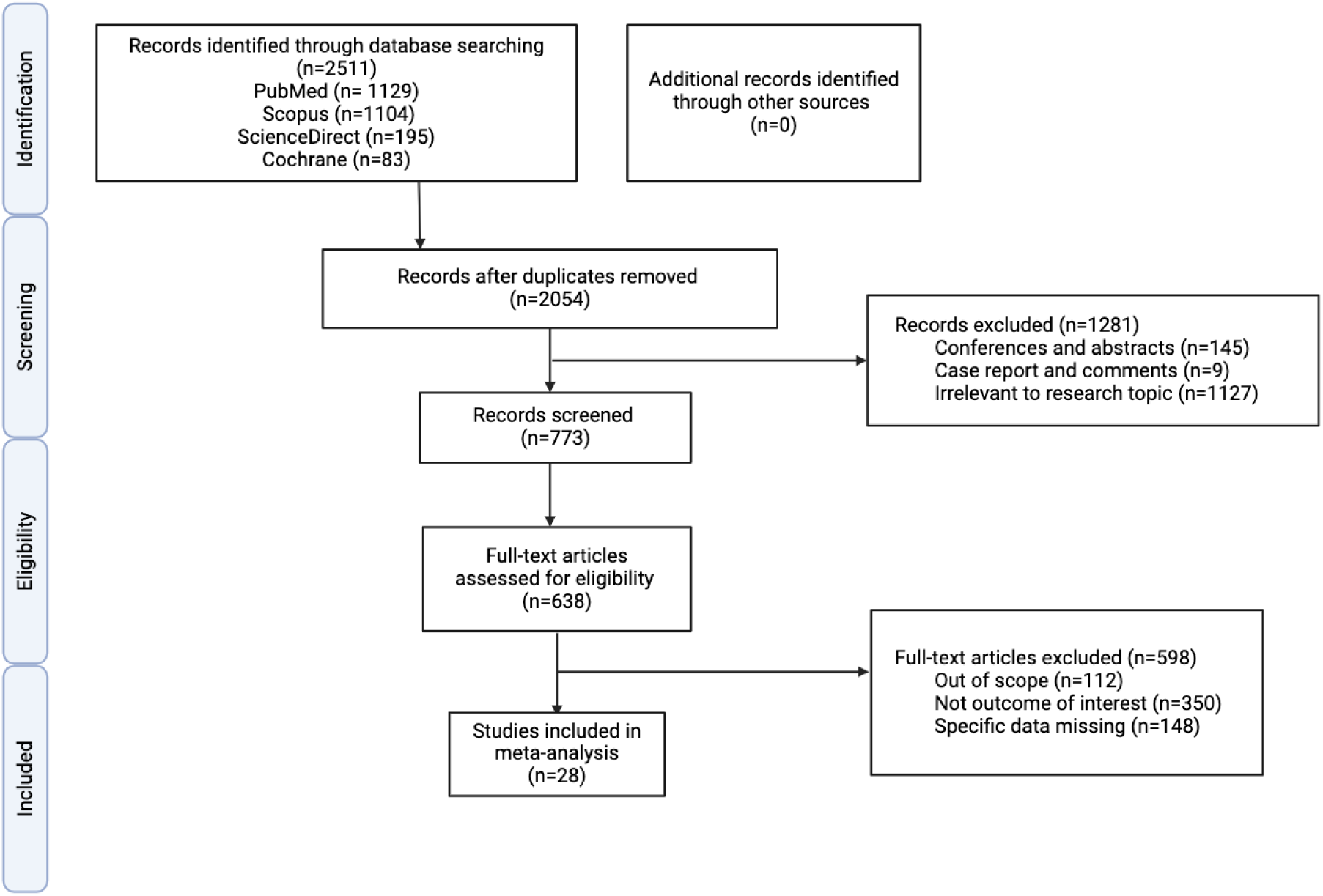
PRISMA flow diagram for the literature search and study selection process. The diagram outlines the workflow across the four stages of study selection: identification, screening, eligibility, and inclusion. It illustrates the number of records retrieved, screened, excluded, and ultimately included in the final analysis, adhering to PRISMA guidelines.

**Table 1:**
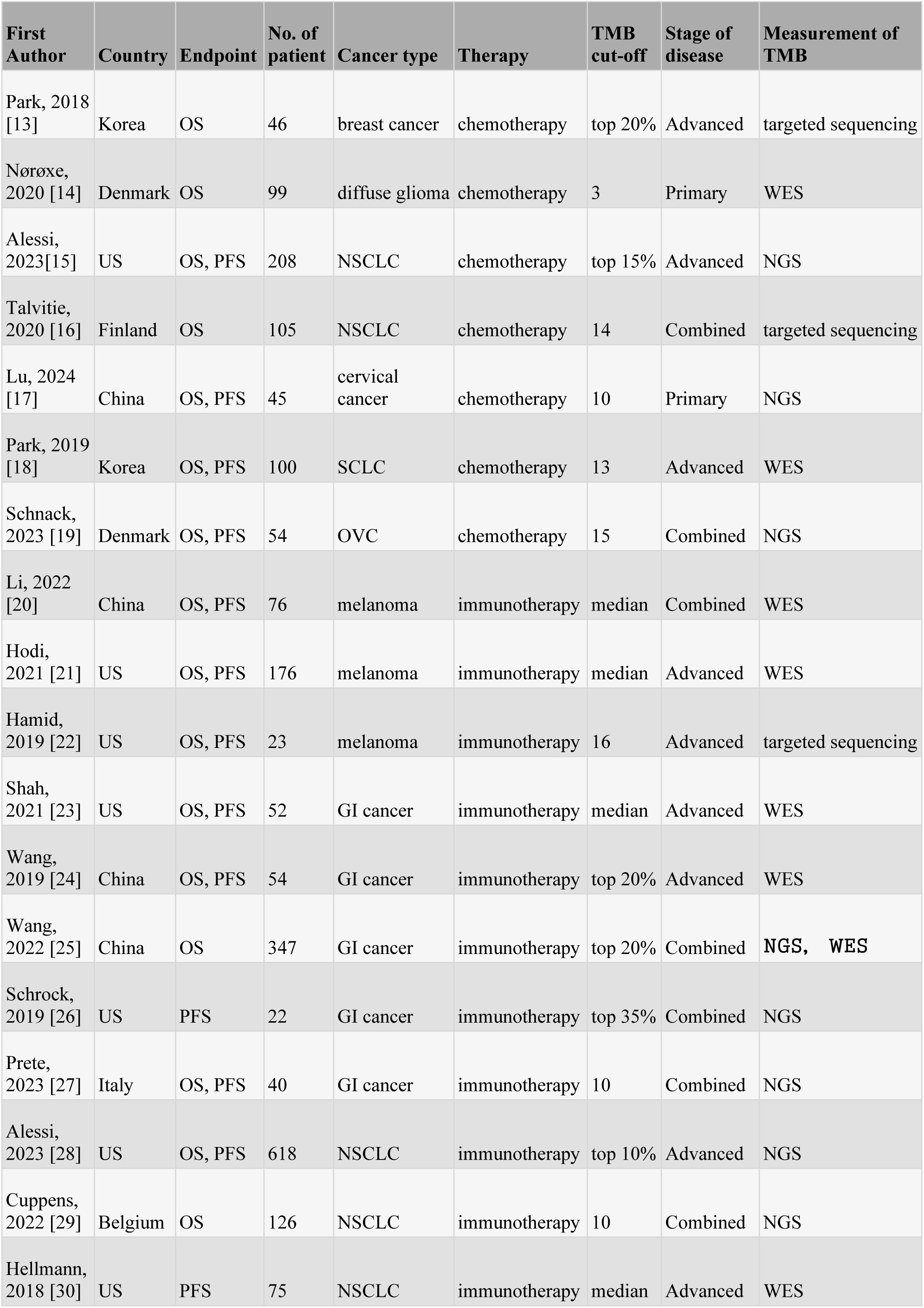
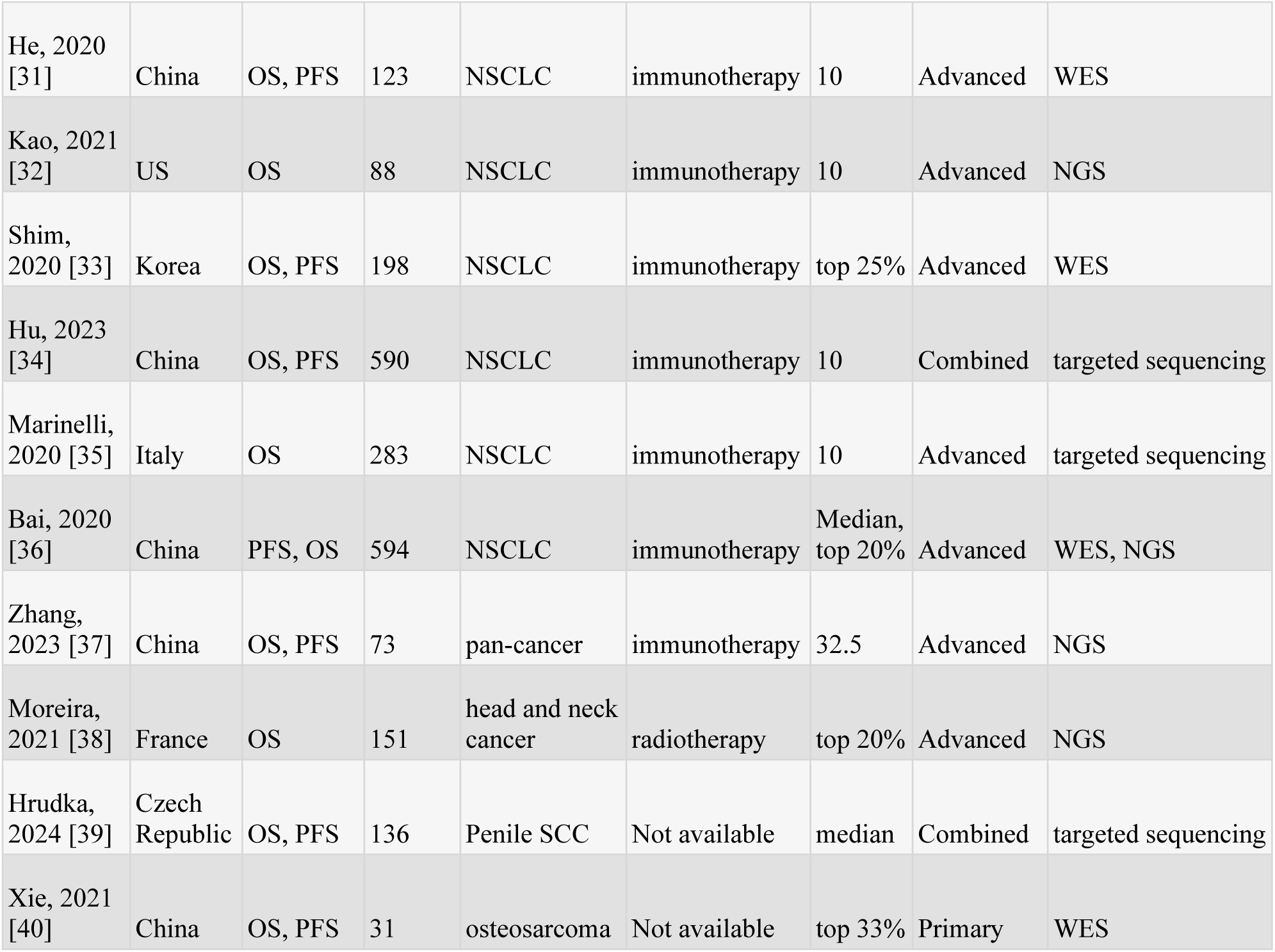
Key characteristics of the studies included in the meta-analysis.

### Meta-analysis of survival outcomes

A meta-analysis of 34 cohorts from 25 studies (4,928 cases) for overall survival (OS) and 25 cohorts from 19 studies (3,349 cases) for progression-free survival (PFS) revealed that high TMB was significantly associated with improved OS and PFS. For OS, the pooled hazard ratio (HR) was 0.53 (95% CI: 0.45 to 0.62, I² = 58%, *p* < 0.01), while for PFS, the HR was 0.56 (95% CI: 0.47 to 0.67, I² = 61%, *p* < 0.01) (**Figure 2A, 2B)**. Moderate between-study heterogeneity was observed for OS and PFS, driven by differences in cancer types, treatment modalities, and TMB thresholds. Notably, a paired *t*-test showed a statistically significant difference between HRs for OS and PFS (*p* = 0.0409) (**Figure 2C**), suggesting that TMB may exert distinct prognostic influences depending on the clinical endpoint. Moreover, some studies had wider CI intervals, reflecting greater uncertainty, potentially due to the small sample sizes and rare incidence. To explore sources of heterogeneity, subgroup analyses were performed based on cancer type, treatment, TMB thresholds, and disease stage.

**Figure 2.**
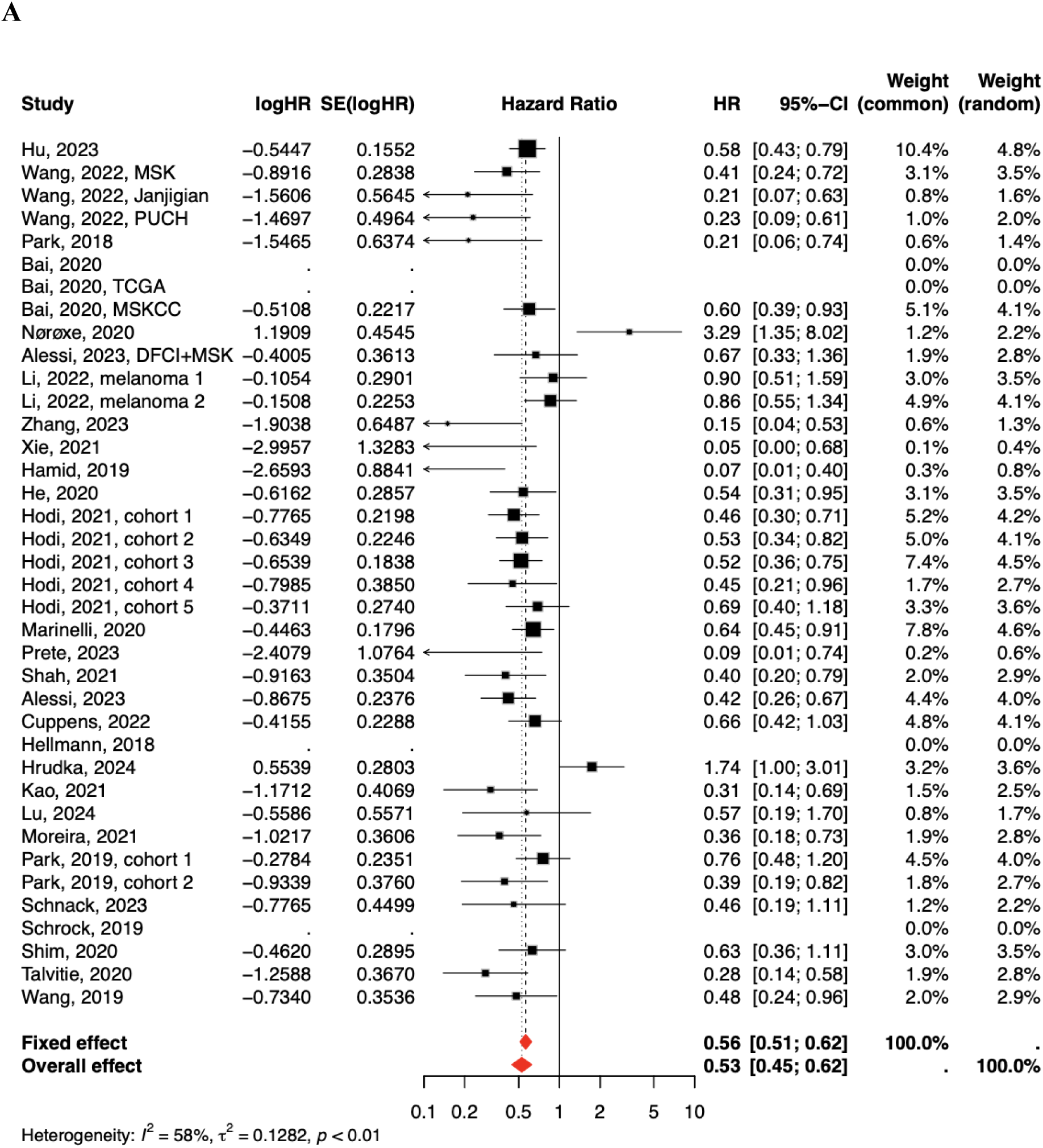
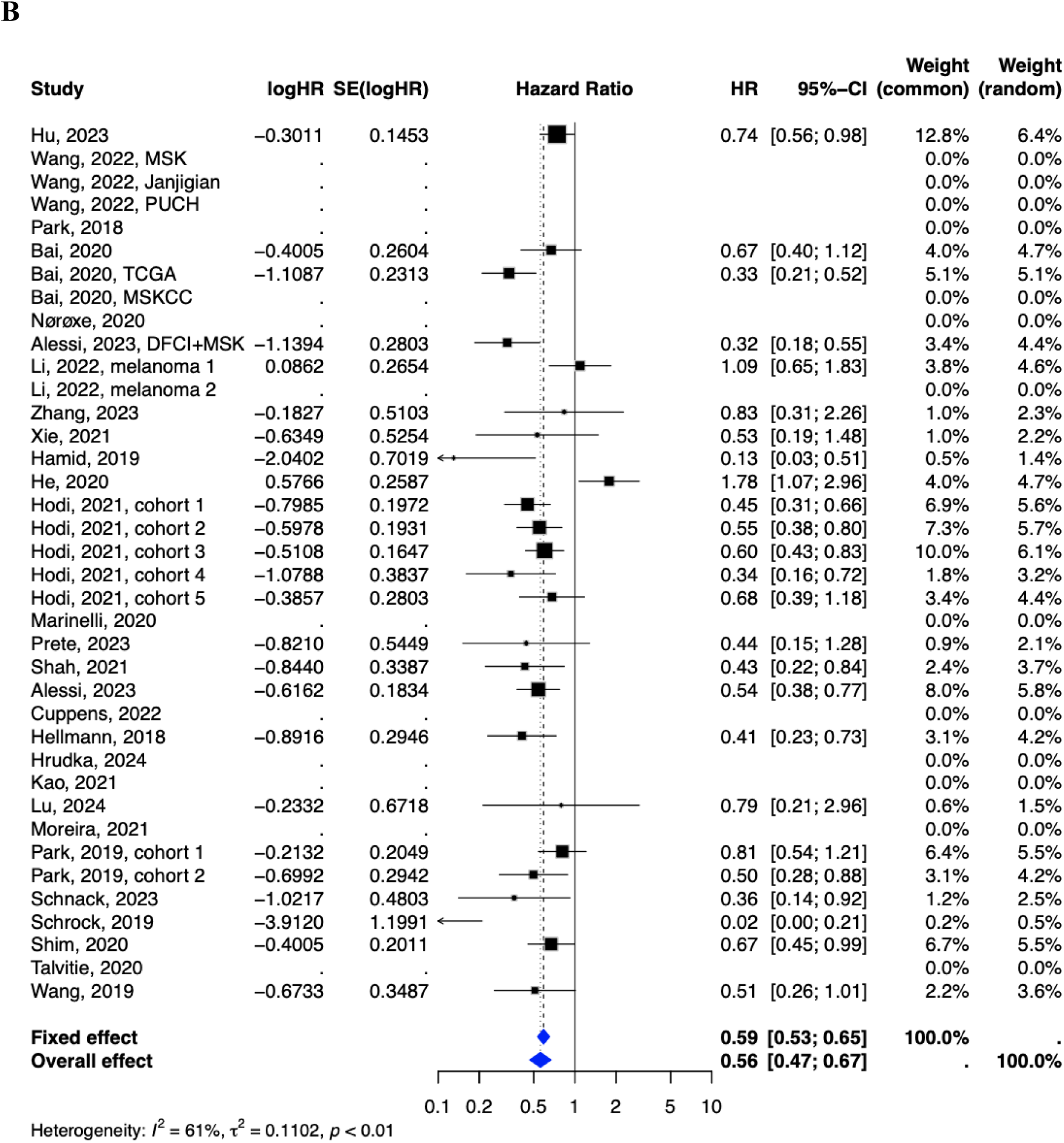
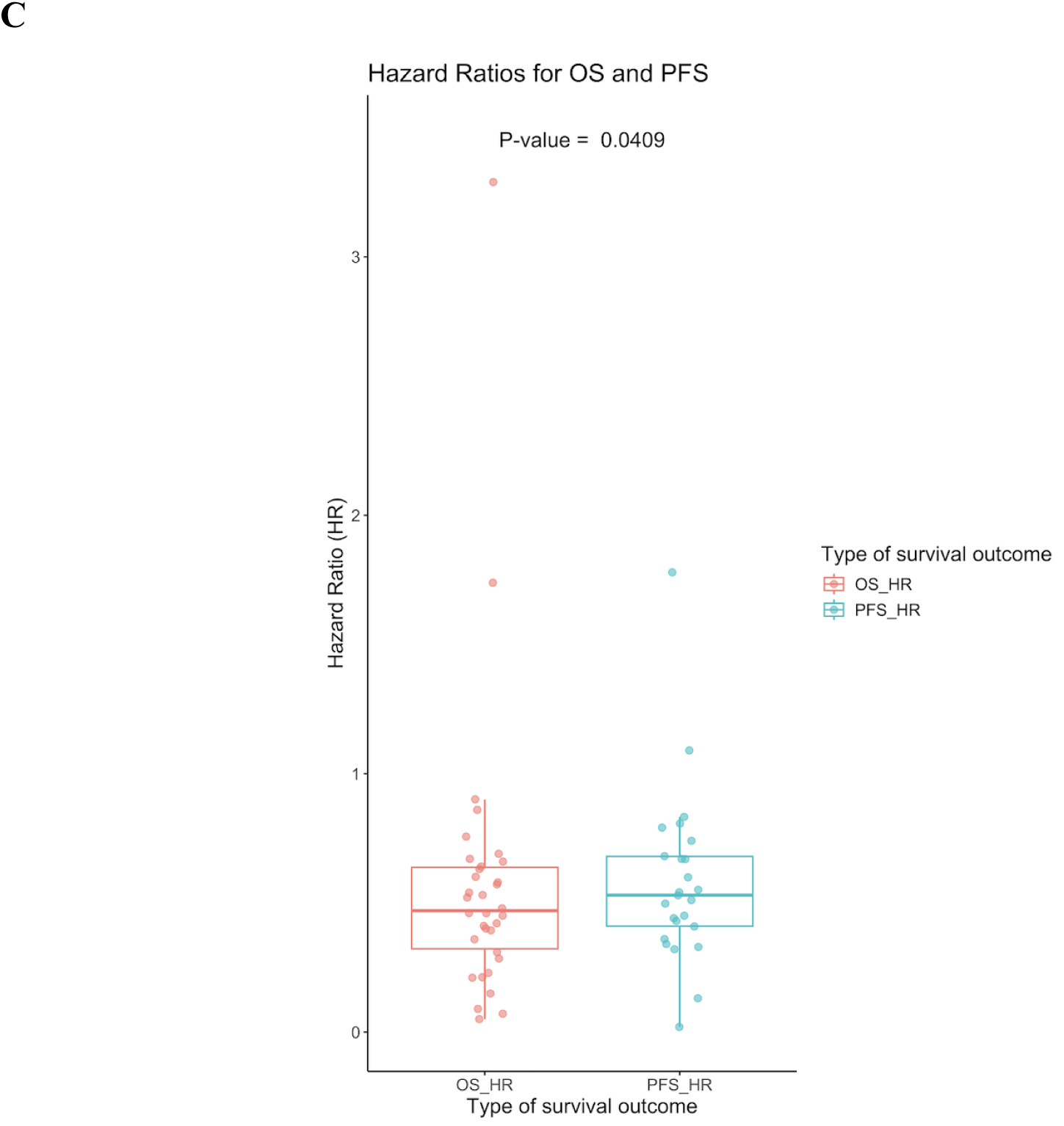
Association between high TMB and survival outcomes. (**A, B**) Forest plots for OS and PFS illustrate the meta-analysis results using random-effects models, showing HRs, 95% CIs, effect sizes, and between-study heterogeneity. (**C**) Boxplot of HRs from included studies for OS and PFS highlights the pooled results, demonstrating statistically significant differences in the impact of high TMB on OS and PFS (*p* = 0.0409). Despite observed between-study heterogeneity, high TMB is consistently associated with longer survival outcomes for both OS and PFS.

### Prognostic impact of TMB across cancer types

High TMB was associated with significantly improved OS and PFS in immunogenic cancers such as NSCLC (OS: HR = 0.56, 95% CI: 0.48 to 0.64, I² = 0%; PFS: HR = 0.59, 95% CI: 0.41 to 0.85, I² = 79%) and melanoma (OS: HR = 0.58, 95% CI: 0.47 to 0.72, I² = 48%; PFS: HR = 0.56, 95% CI: 0.42 to 0.74, I² = 58%) (**Figure 3A, 3B**). These cancers are characterised by a high neoantigen load, which enhances immune system recognition and increases the efficacy of immune checkpoint inhibitors (ICIs). Similarly, GI cancers demonstrated higher survival (OS: HR = 0.36, 95% CI: 0.26 to 0.50, I² = 0%; PFS: HR = 0.42, 95% CI: 0.27 to 0.64, I² = 56%), consistent with hypermutated subtypes showing increased sensitivity to ICIs.

**Figure 3.**
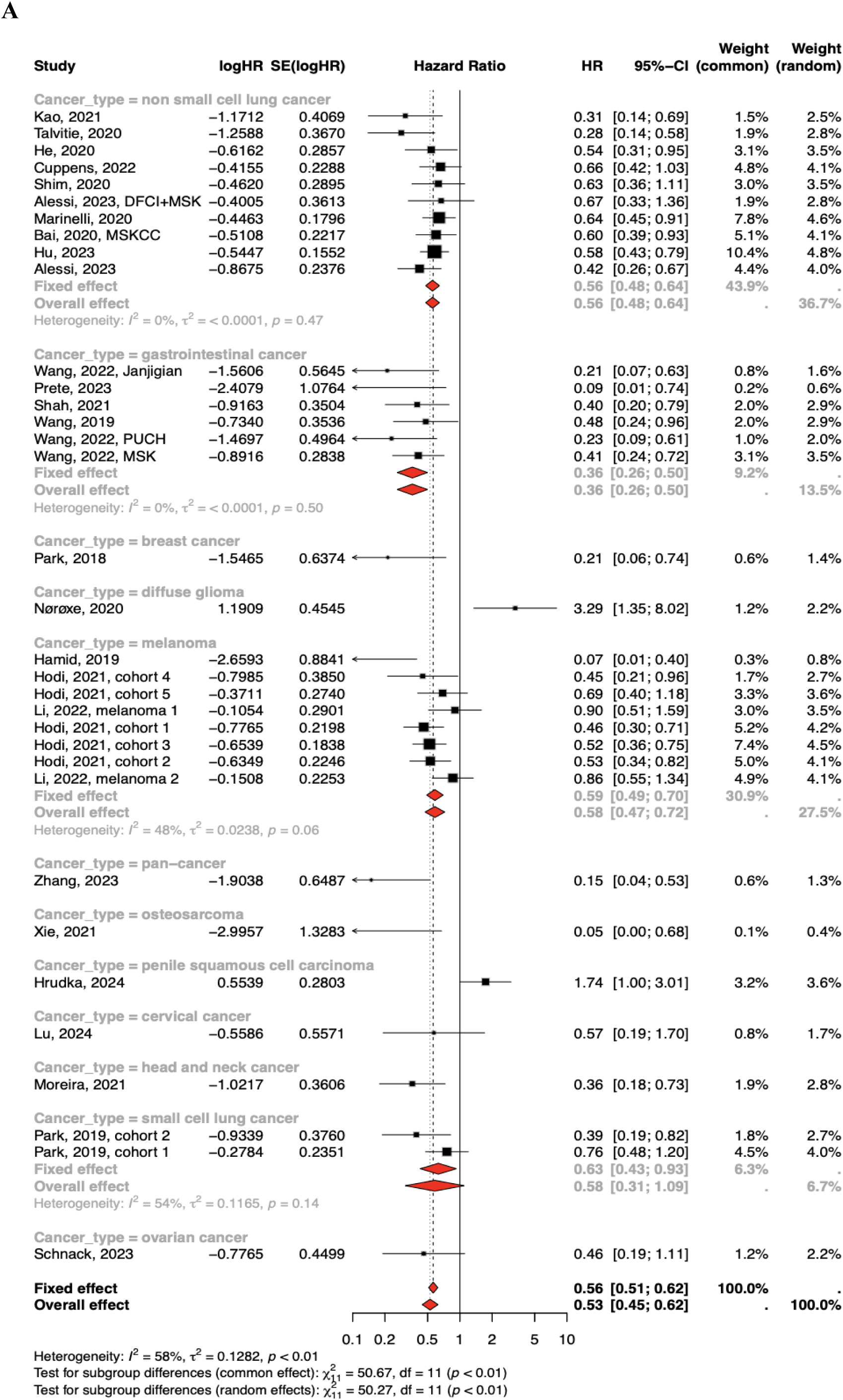
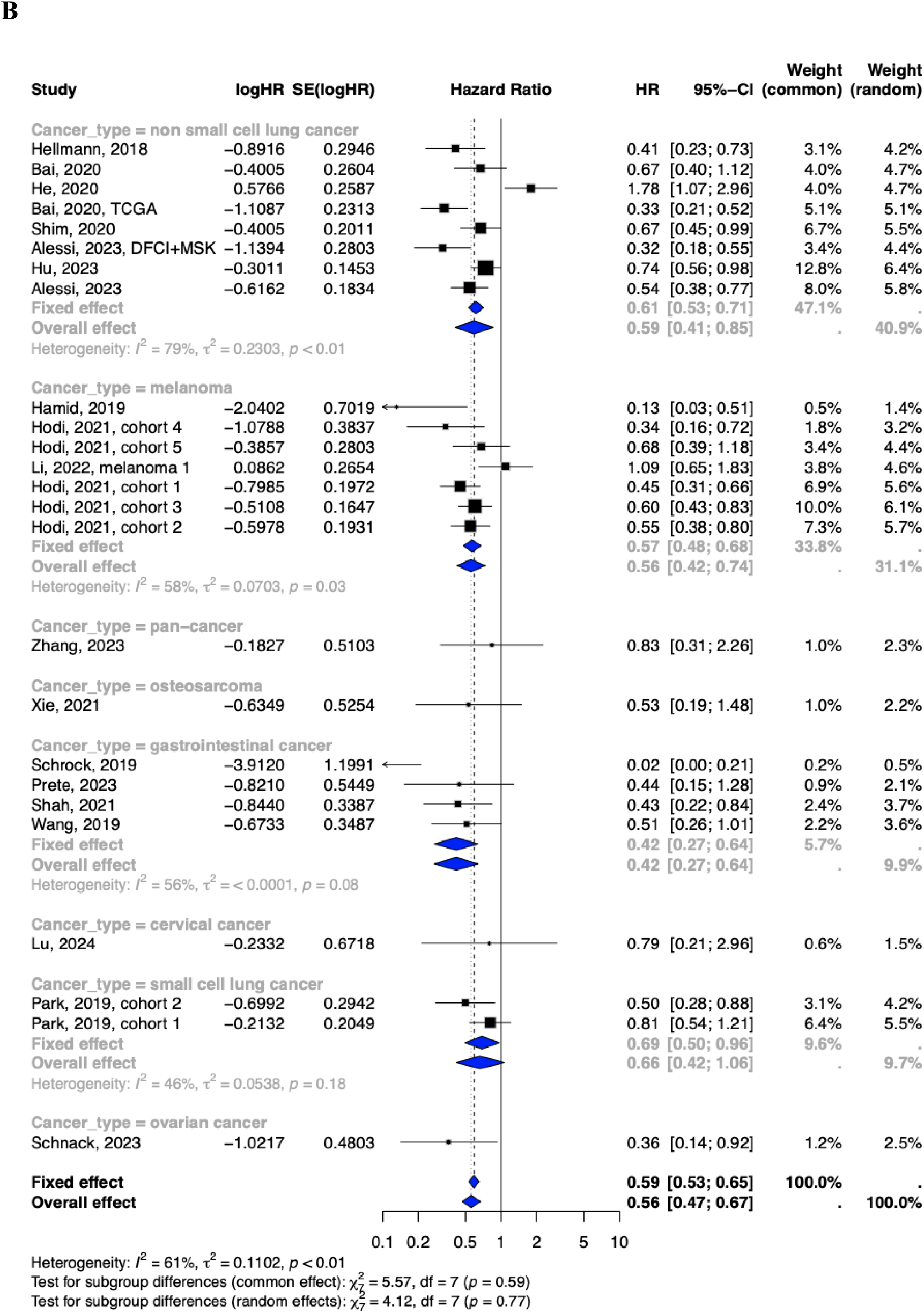
Subgroup analyses of the prognostic value of high tumour mutation burden (TMB) in patients stratified by cancer type for **A)** overall survival (OS) and **B)** progression-free survival (PFS). High TMB was associated with improved OS and PFS across most cancer types. Subgroup analyses revealed low to moderate between-study heterogeneity for OS and low to high heterogeneity for PFS.

In contrast, gliomas and penile SCC exhibited worse OS with high TMB (gliomas: HR = 3.29, 95% CI: 1.35 to 8.02; penile SCC: HR = 1.74, 95% CI: 1.00 to 3.01). Gliomas, known for their immunosuppressive microenvironment dominated by regulatory T cells, may undermine the therapeutic benefit of high TMB. Similarly, penile SCC, often associated with human papillomavirus (HPV) infection, may exhibit tumour-immune interactions that reduce the predictive value of TMB for survival outcomes. Notably, PFS data for gliomas and penile SCC were unavailable, likely reflecting the limited representation of these cancers in the included studies.

These findings highlight the cancer-specific nature of TMB’s prognostic value and underscore the importance of accounting for tumour biology and immune microenvironments in interpreting the moderate heterogeneity observed in OS (I² = 58%) and PFS (I² = 61%) analyses. Subgroup analyses revealed that heterogeneity was lower in immunogenic cancers (e.g., NSCLC, melanoma) where TMB thresholds and sequencing platforms were more standardised. Variability in immune microenvironments, treatment regimens, and TMB cut-offs likely contributed to differences in survival outcomes across studies. These findings suggest that refining TMB measurement methods and developing cancer-type-specific thresholds are essential to improving its clinical utility as a biomarker.

### Influence of treatment modality on TMB prognostic value

We evaluated whether treatment type contributed to heterogeneity in survival outcomes (**Figure 4**). The results indicate that immunotherapy-treated cohorts with high TMB consistently display better OS compared to PFS, with lower between-study heterogeneity for OS (HR = 0.53, 95% CI: 0.48 to 0.60, I² = 37%, *p* = 0.05, **Figure 4A**) than for PFS (HR = 0.56, 95% CI: 0.45 to 0.70, I² = 68%, *p* < 0.01, **Figure 4B**). These findings confirm the stronger predictive value of TMB for overall survival in the context of immunotherapy. In chemotherapy-treated cohorts, the association of high TMB with survival outcomes is less consistent. High TMB is associated with better OS (HR = 0.60, 95% CI: 0.38 to 0.95, I² = 66%, *p* < 0.01) and longer PFS (HR = 0.55, 95% CI: 0.39 to 0.77, I² = 44%, *p* = 0.11). These results demonstrate that TMB may be a less reliable biomarker in chemotherapy-treated cohorts, reflecting the different mechanisms of action between chemotherapy and immunotherapy.

**Figure 4.**
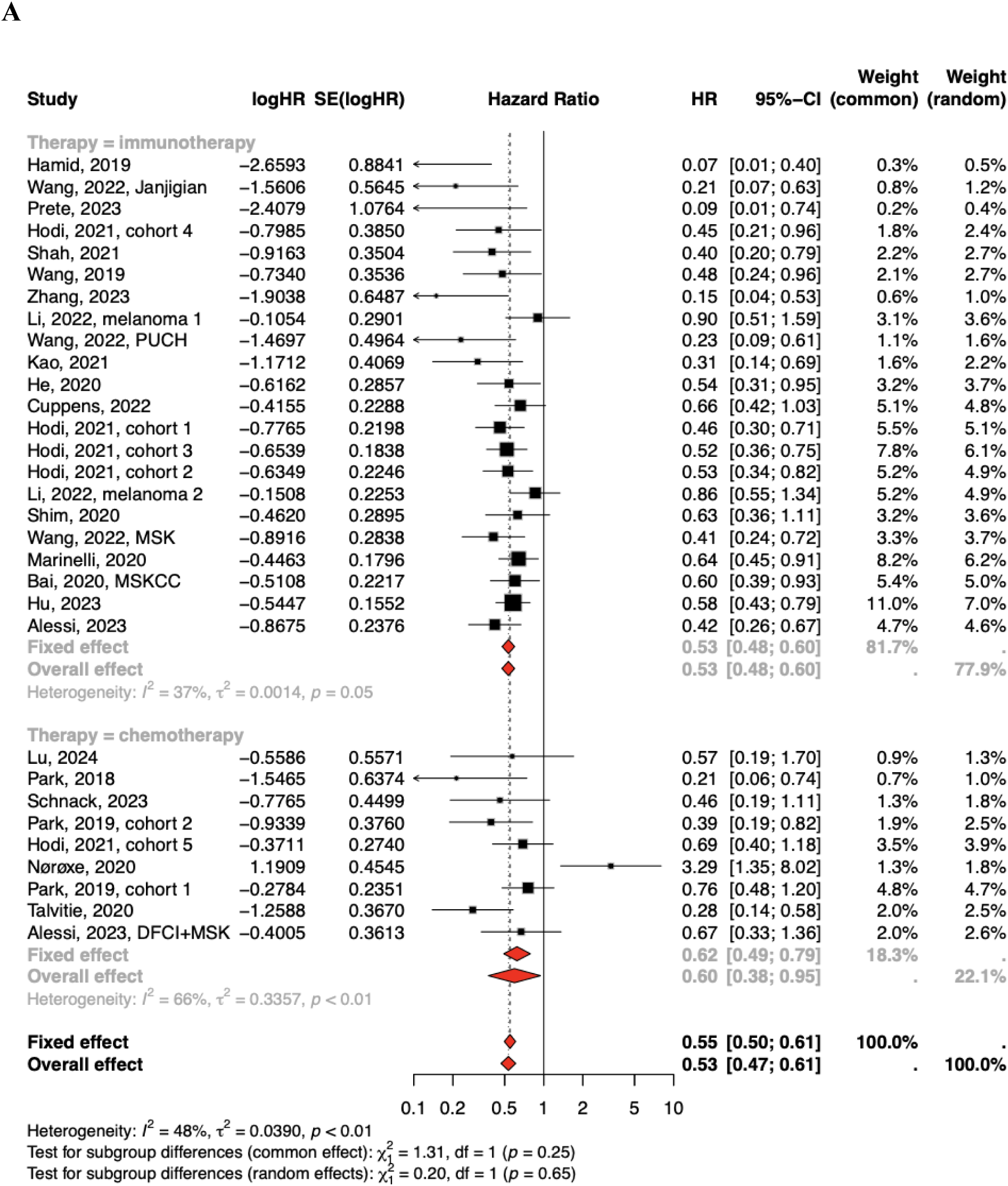
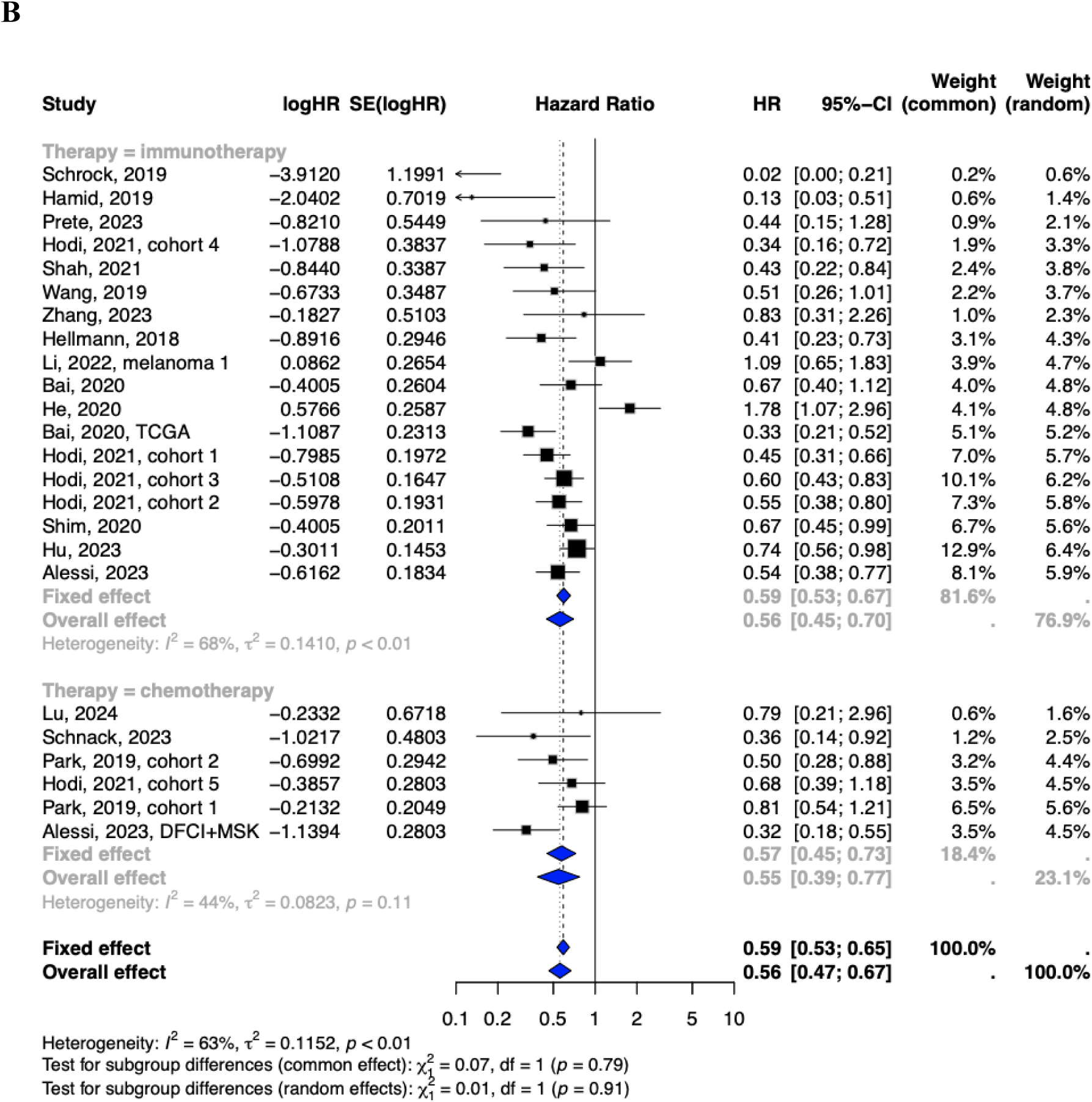
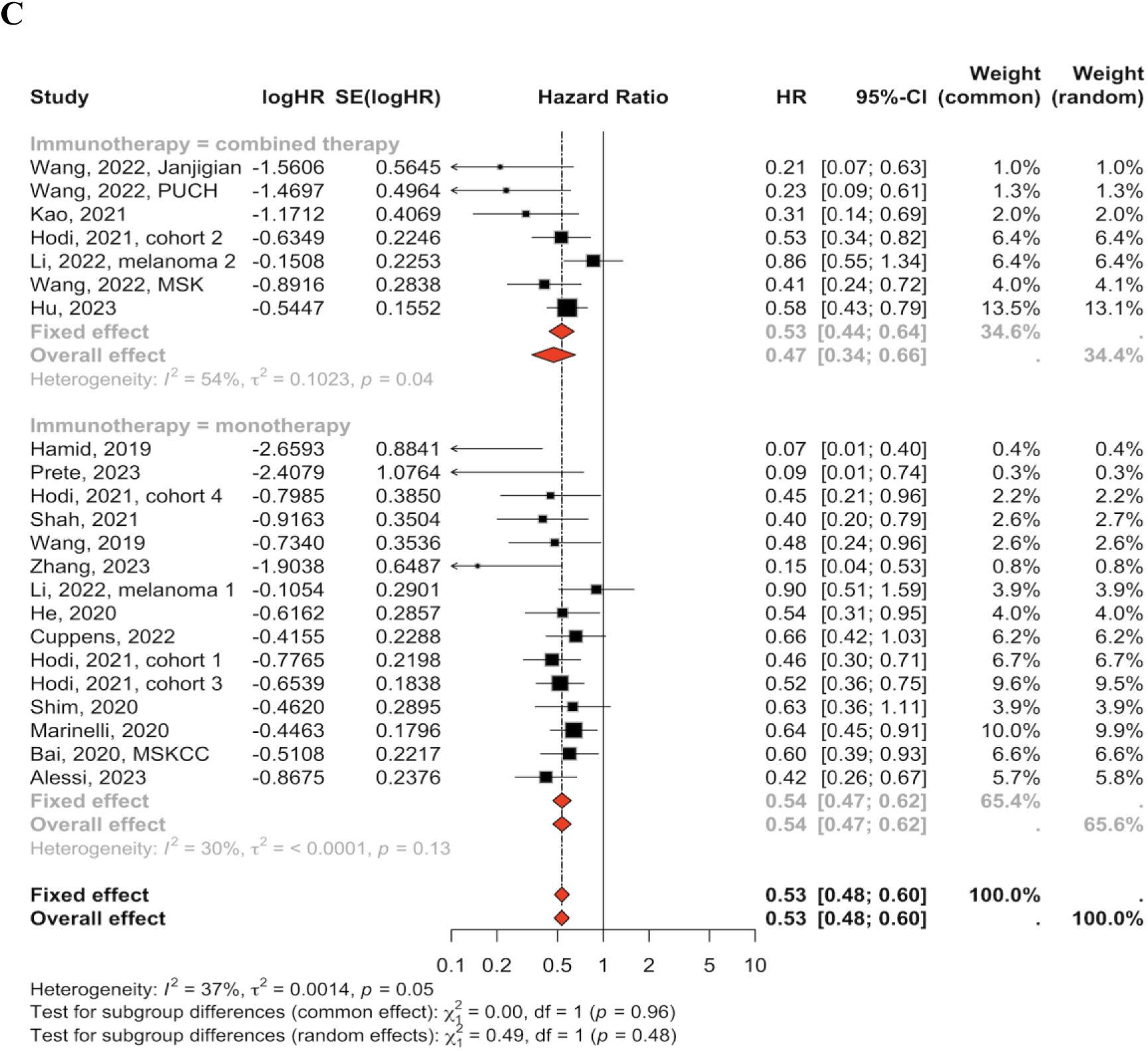
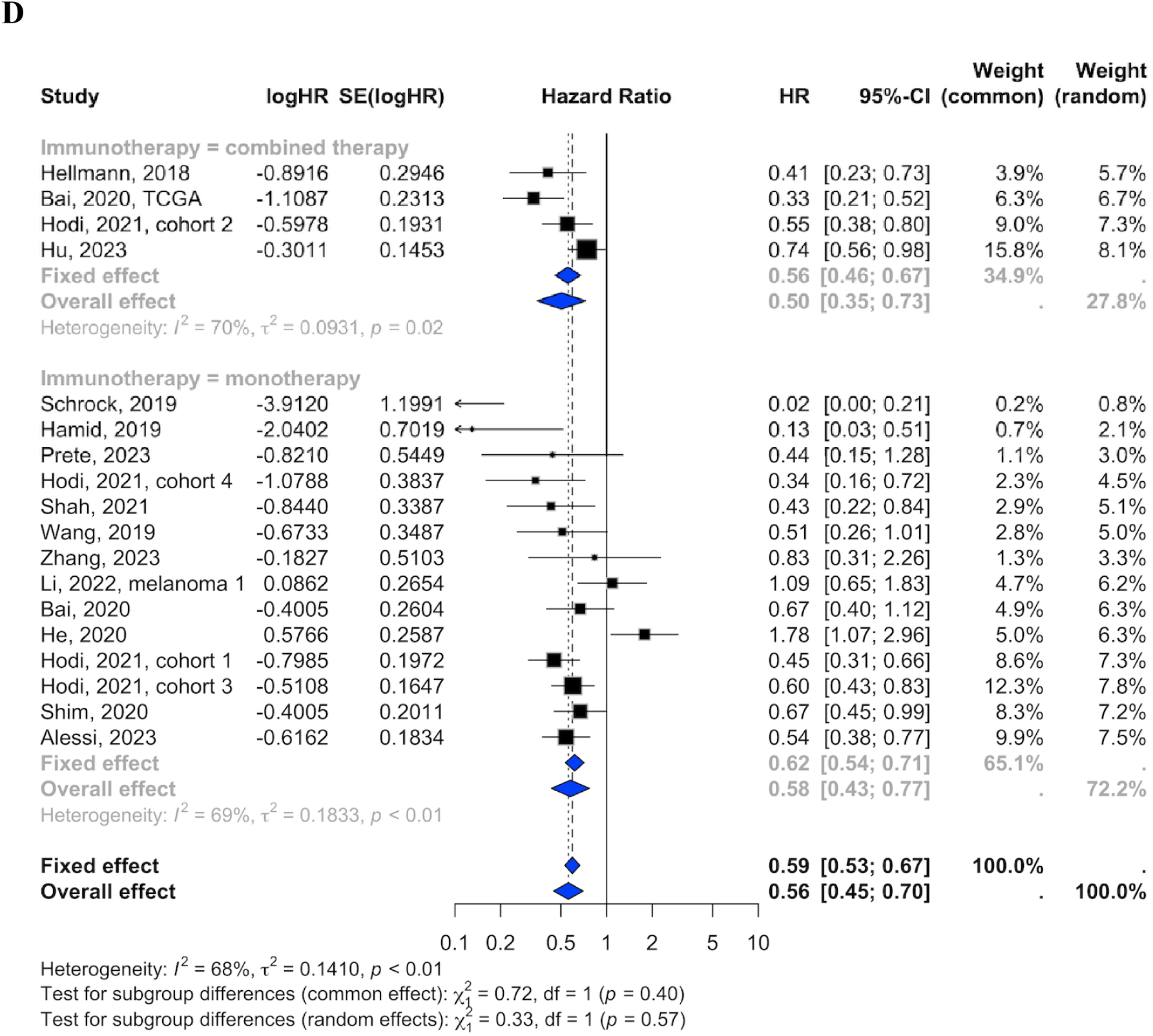
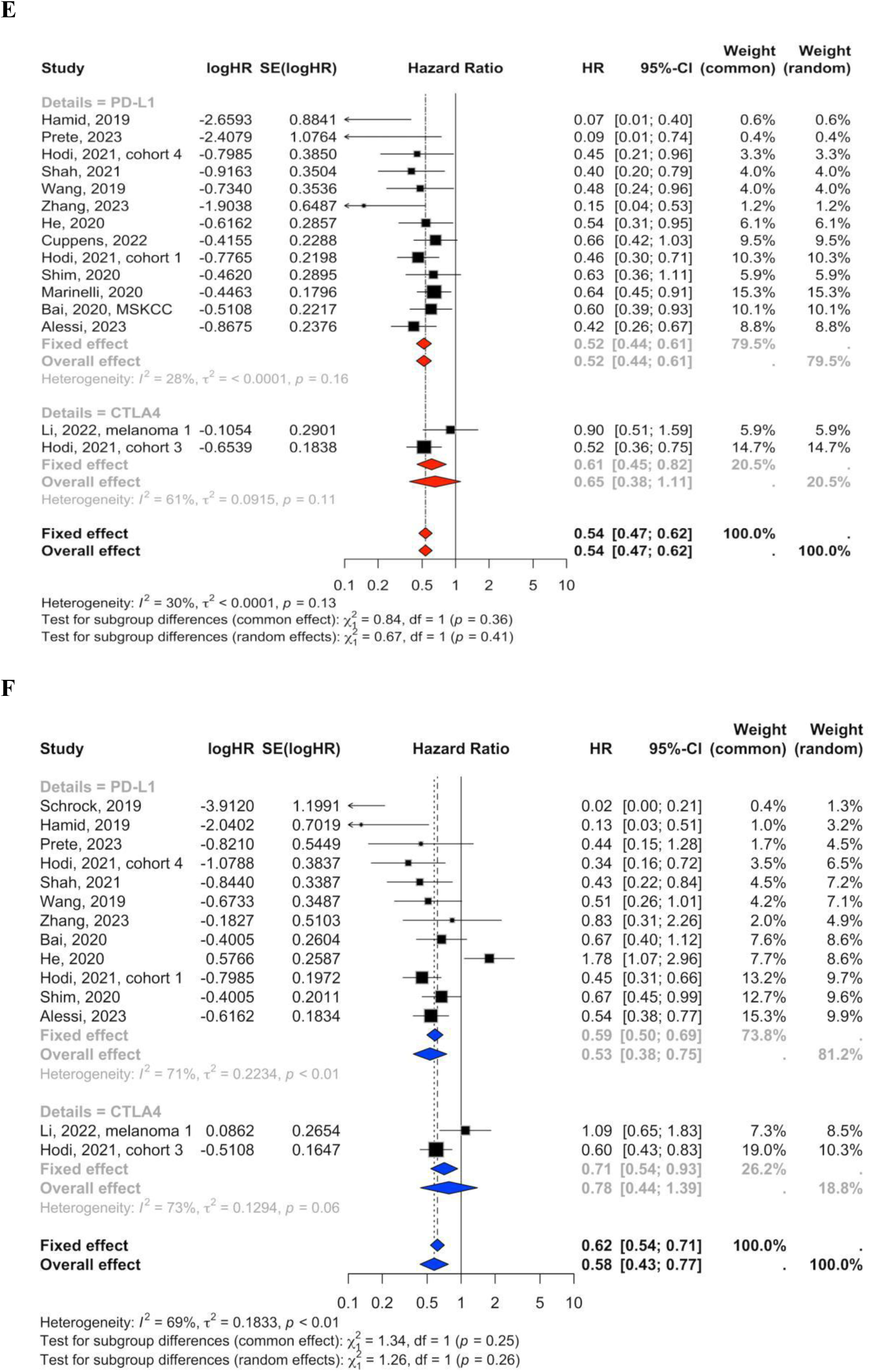
Subgroup analyses of the association between high tumour mutation burden (TMB) and survival outcomes by treatment type. (**A, B**) High TMB is associated with better overall survival (OS) and progression-free survival (PFS) in immunotherapy-treated cohorts, with lower between-study heterogeneity for OS compared to PFS. (**C, D**) Subgroup analyses within immune checkpoint inhibitor (ICI) therapies show a stronger association with improved OS for anti-PD-L1/PD-1 treatments than anti-CTLA-4 therapies, while PFS benefits remain inconsistent in mono-immunotherapy cohorts. (**E, F**) Combination therapy using both anti-PD-L1/PD-1 and anti-CTLA-4 inhibitors demonstrates the strongest association with improved OS and PFS outcomes, reflecting a synergistic effect in dual immune checkpoint blockade. These results highlight the role of TMB as a predictive biomarker, particularly in immunotherapy-treated patients.

Within ICI therapies, high TMB is more strongly associated with better survival outcomes in cohorts treated with combined therapies (for OS, HR = 0.53, 95% CI: 0.44 to 0.61, I² = 54%, *p* = 0.04, **Figure 4C**; for PFS, HR = 0.50, 95% CI: 0.35 to 0.73, I² = 70%, *p* = 0.02, **Figure 4D**) compared to monotherapy (for OS, HR = 0.54, 95% CI: 0.47 to 0.62, I² = 30%, *p* = 0.13; for PFS, HR = 0.58, 95% CI: 0.43 to 0.77, I² = 69%, *p* < 0.01), highlighting a synergistic effect. These findings align with the established efficacy of ICI therapies in diverse tumour types.

Further stratification by ICI class revealed superior survival outcomes in high-TMB patients treated with anti-PD-L1/PD-1 inhibitors compared to anti-CTLA4 therapy. Among patients receiving anti-PD-L1/PD-1 monotherapy, high TMB was associated with a significant improvement in OS (HR = 0.52, 95% CI: 0.44–0.61, I² = 28%, p = 0.16, **Figure 4E**) and PFS (HR = 0.53, 95% CI: 0.38–0.75, I² = 71%, p < 0.01, **Figure 4F**). In contrast, the anti-CTLA4 monotherapy cohort exhibited weaker and more variable survival benefits (OS: HR = 0.65, 95% CI: 0.38–1.11, I² = 61%, p = 0.11; PFS: HR = 0.78, 95% CI: 0.44–1.39, I² = 73%, p = 0.06), suggesting differences in tumour-intrinsic sensitivity to immune modulation.

Due to the limited number of studies evaluating radiotherapy-treated cohorts, this subgroup was excluded from the analysis. The small sample size lacked sufficient statistical power to provide reliable conclusions. Further research is necessary to investigate the predictive role of TMB in radiotherapy outcomes.

These findings establish TMB as a predictive biomarker for immune checkpoint inhibitor (ICI) therapies, particularly in identifying patients who benefit from anti-PD-L1/PD-1 therapies and combination immunotherapy strategies. The enhanced survival outcomes in high TMB cohorts are driven by increased neoantigen presentation, which activates immune responses and promotes tumour clearance. In chemotherapy-treated patients, the predictive value of TMB is less reliable, likely reflecting the reliance of chemotherapy on cytotoxic mechanisms rather than immune activation. Combination therapies targeting multiple immune checkpoints yield the strongest survival benefits, indicating that high TMB amplifies the efficacy of dual immune modulation. These results provide a robust foundation for tailoring treatment strategies based on TMB levels to optimise therapeutic outcomes and improve survival in patients with solid tumours.

### Defining optimal TMB thresholds for prognostic stratification

TMB cut-offs are crucial for stratifying patients to identify subgroups with distinct therapeutic responses and prognostic outcomes, yet no universally accepted standard exists. In this study, we applied two commonly used thresholds: a fixed value of 10 mut/Mb and the top 20% of TMB values within cohorts. Both cut-offs consistently demonstrated a strong association between high TMB and improved survival outcomes. For OS, the HR was 0.58 (95% CI: 0.48 to 0.69, I² = 1%, *p* = 0.42) using the 10 mut/Mb cut-off and 0.44 (95% CI: 0.36 to 0.55, I² = 6%, *p* = 0.39) for the top 20% cut-off (**Figure 5A**). For PFS, the HR was 0.90 (95% CI: 0.49 to 1.64, I² = 71%, *p* = 0.01) with the 10 mut/Mb cut-off and 0.46 (95% CI: 0.33 to 0.64, I² = 20%, *p* = 0.29) for the top 20% cut-off (**Figure 5B**). These findings suggest that higher somatic mutation loads, reflected by high TMB, enhance tumour immunogenicity by increasing the production of neoantigens. These neoantigens, recognised by the immune system, can boost the efficacy of immune checkpoint inhibitors (ICIs), leading to prolonged and durable responses. The differences elicited based on the two cut-offs also underscore the importance of selecting biologically and clinically relevant thresholds. Notably, the top 20% cut-off showed lower heterogeneity and stronger associations, suggesting it may better capture tumour-specific variations in mutation load.

**Figure 5.**
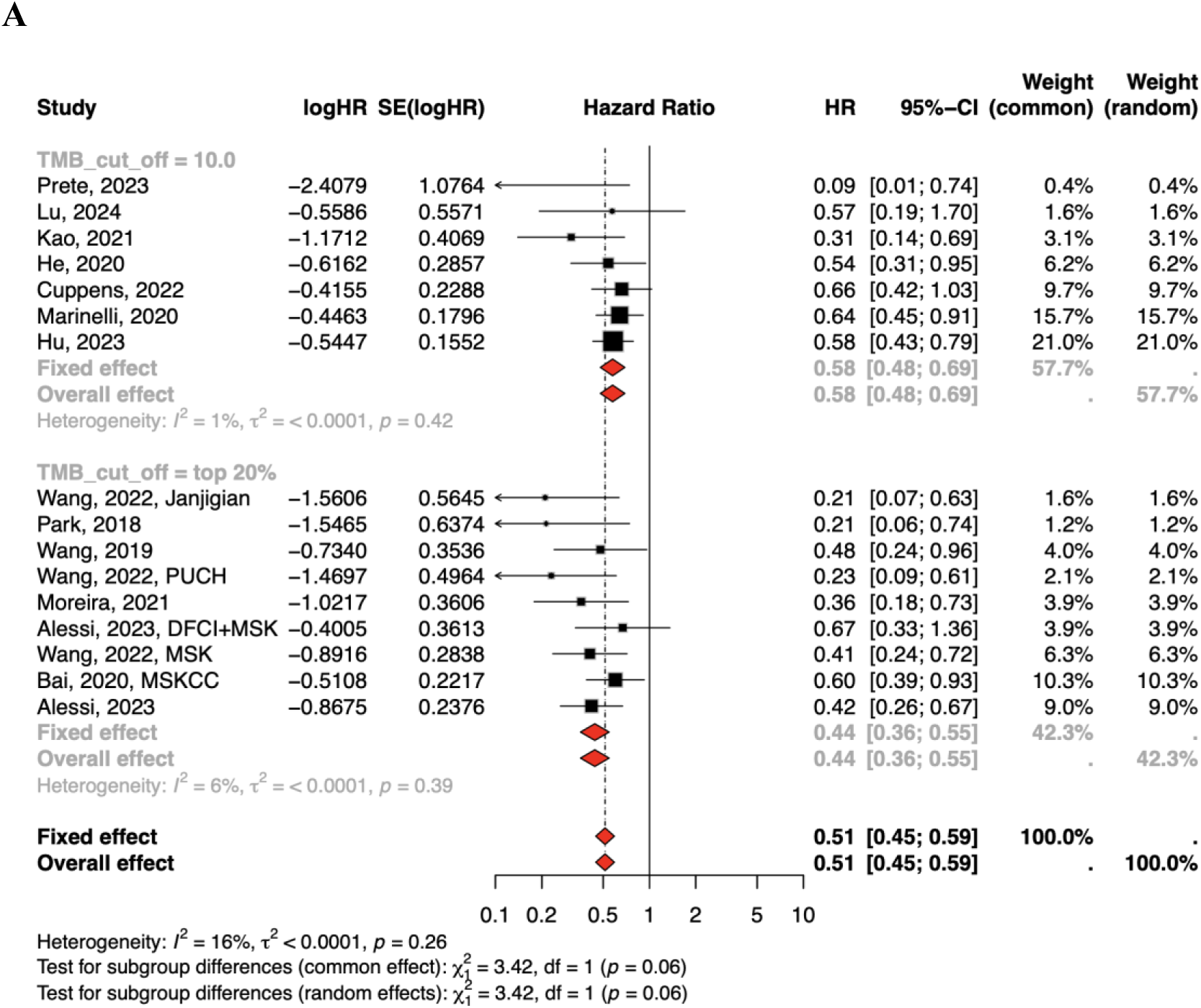
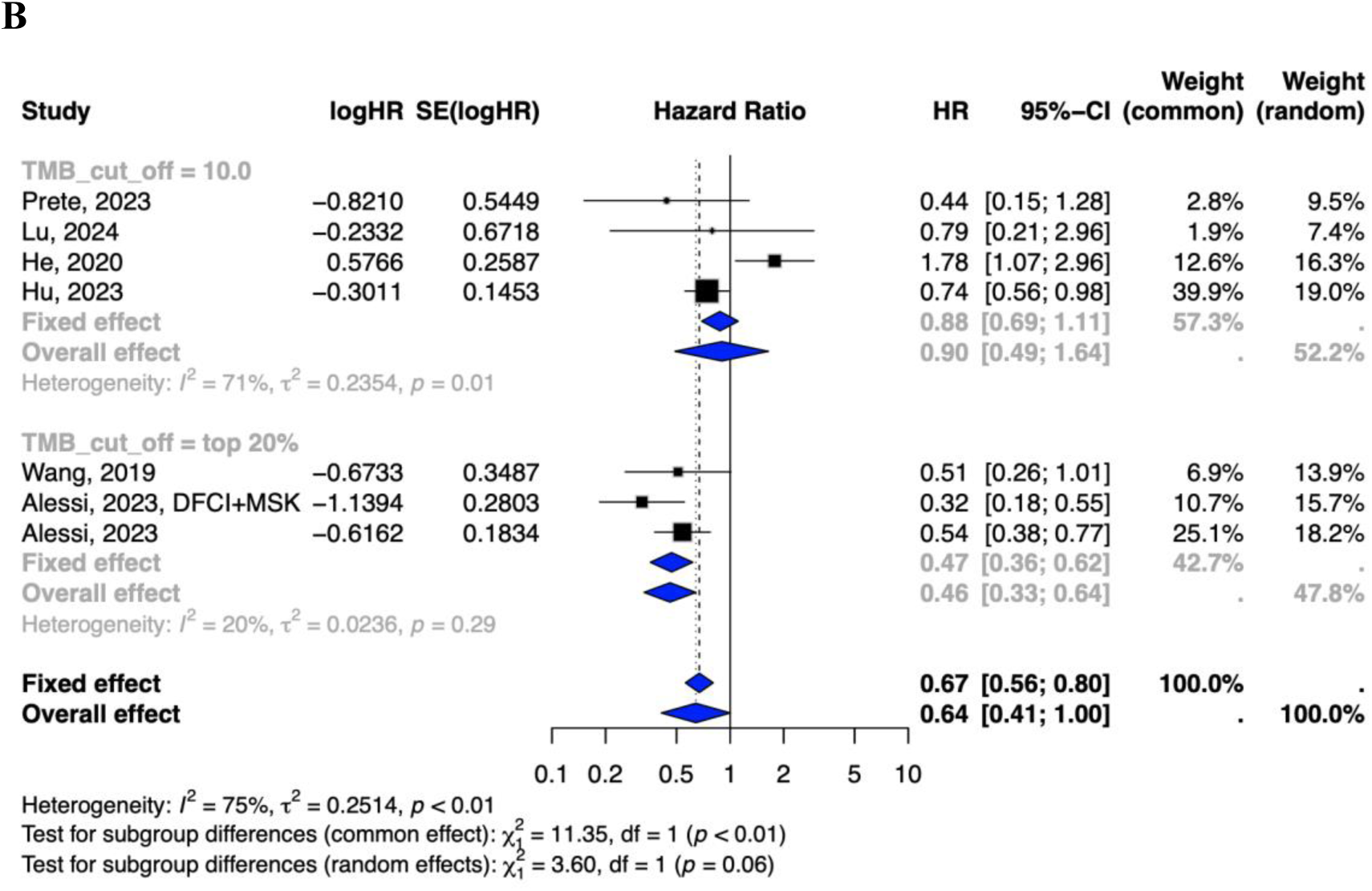
Subgroup analyses of survival outcomes by TMB cut-off values of 10 mut/Mb and the top 20%. (**A**) Overall survival (OS) and (**B**) progression-free survival (PFS) analyses show the predictive value of high TMB for improved survival outcomes using both cut-offs. Stratification by the top 20% cut-off demonstrates stronger associations and lower between-study heterogeneity than the fixed 10 mut/Mb threshold.

### Disease stage-specific prognostic implications of TMB

Tumour progression and disease stage significantly influence survival outcomes and may alter the predictive value of TMB. Subgroup analyses revealed that high TMB was associated with better OS and PFS outcomes in both primary and advanced/recurrent disease stages (**Figure 6**).

**Figure 6.**
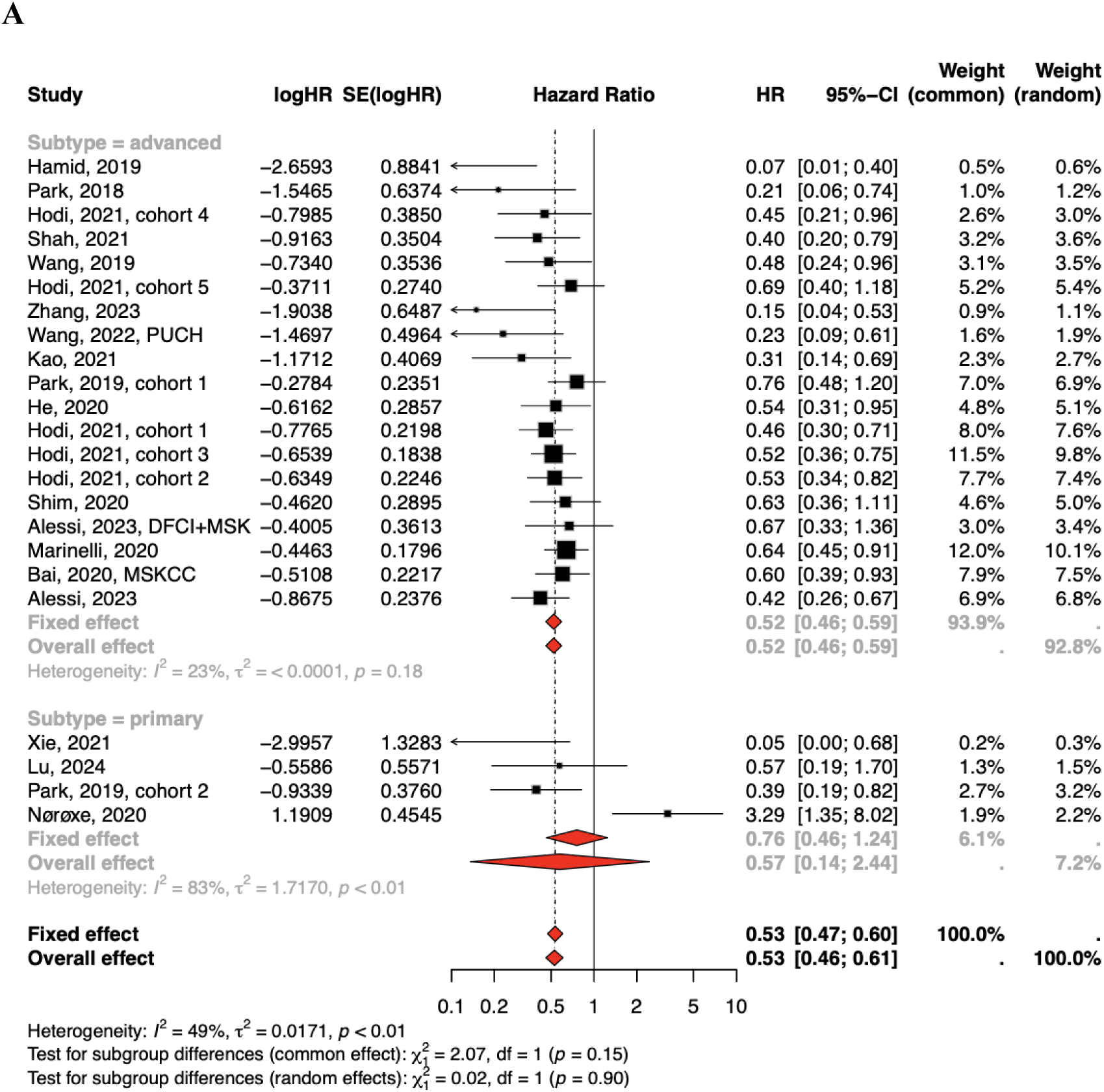
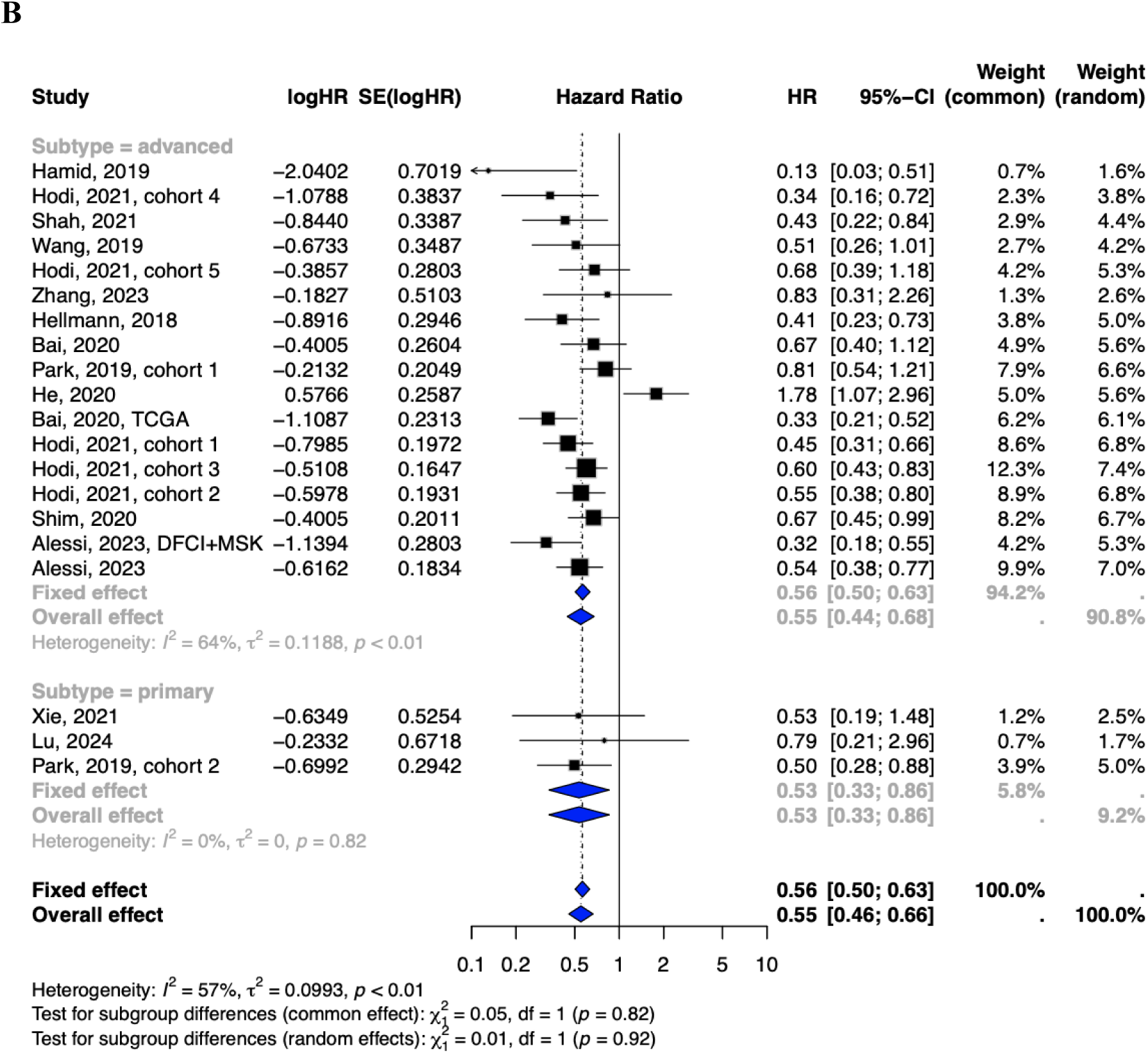
Subgroup analyses of survival outcomes by disease stage. (**A**) Overall survival (OS) and (**B**) progression-free survival (PFS) analyses show that high TMB is associated with improved survival across primary, advanced, and recurrent disease stages. Despite this consistent association, advanced-stage PFS exhibits higher between-study heterogeneity, reflecting the complex interplay of factors influencing survival outcomes in progressive tumours.

In primary stages, high TMB demonstrated a strong association with better survival outcomes (OS: HR = 0.57, 95% CI: 0.14 to 2.44, I² = 83%, *p* < 0.01; PFS: HR = 0.53, 95% CI: 0.33 to 0.86, I² = 0%, *p* = 0.82). Similarly, advanced or recurrent tumours showed improved survival with high TMB (OS: HR = 0.52, 95% CI: 0.46 to 0.59, I² = 23%, *p* = 0.18; PFS: HR = 0.55, 95% CI: 0.44 to 0.68, I² = 64%, *p* < 0.01).

Interestingly, higher heterogeneity was observed in advanced stages compared to primary stages, potentially reflecting the broader diversity of antigen load and the complexity of disease progression in advanced disease. Primary tumours may have a more localised antigen landscape and an intact immune microenvironment, allowing for effective immune surveillance and neoantigen recognition associated with higher TMB. In contrast, advanced or recurrent tumours are often characterised by higher tumour burden and the cumulative effects of prior treatments, which can reshape the tumour-immune interactions. Additionally, factors such as patient fitness and the immunosuppressive effects of advanced disease may further contribute to diminished immune responses, even in the presence of high TMB. These findings highlight TMB as a stage-specific predictive biomarker and the need for further investigation into how disease extent and patient-specific factors influence its prognostic value.

### Impact of sequencing platforms on TMB-based prognostic assessment

The choice of sequencing platforms significantly affects TMB measurement, contributing to variability in survival outcomes (**Figure 7A, 7B**). Commercial platforms such as IMPACT, Foundation Medicine, MSKCC, and Illumina vary in gene coverage, sequencing depth, sensitivity, and mutation detection capabilities, influencing TMB quantification and study comparability. For OS, heterogeneity varied among platforms (I² = 47%, 61%, 58%, and 65%, respectively). For PFS, Illumina and Foundation Medicine platforms showed moderate to high heterogeneity (I² = 54% and 72%, respectively). Despite these discrepancies, high TMB consistently correlated with better OS and PFS outcomes across all sequencing protocols, reaffirming its value as a predictive biomarker. These results underscore the importance of standardising sequencing methods and adopting high-coverage approaches, such as NGS, to improve reproducibility and reliability in TMB assessment.

**Figure 7.**
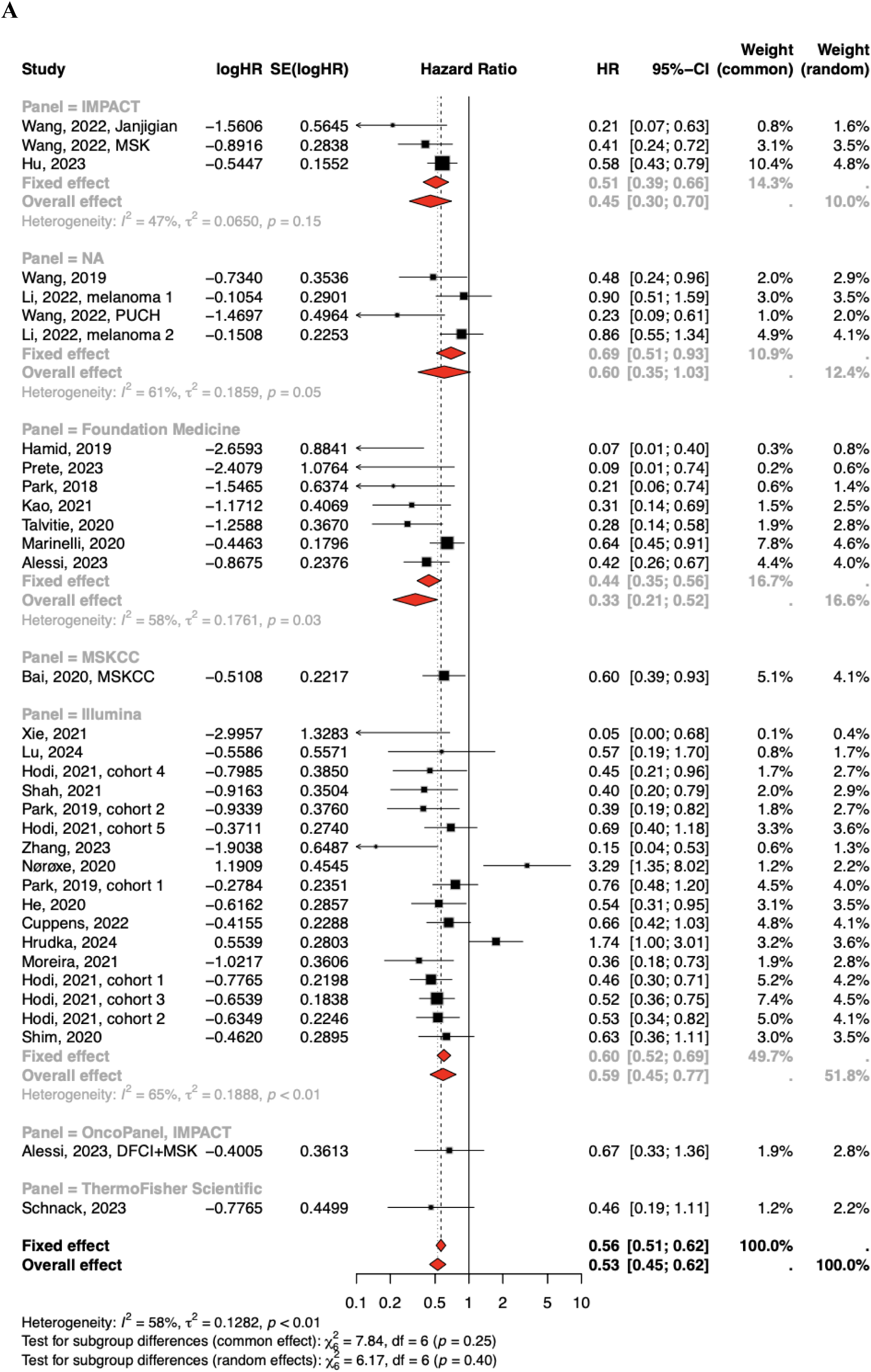
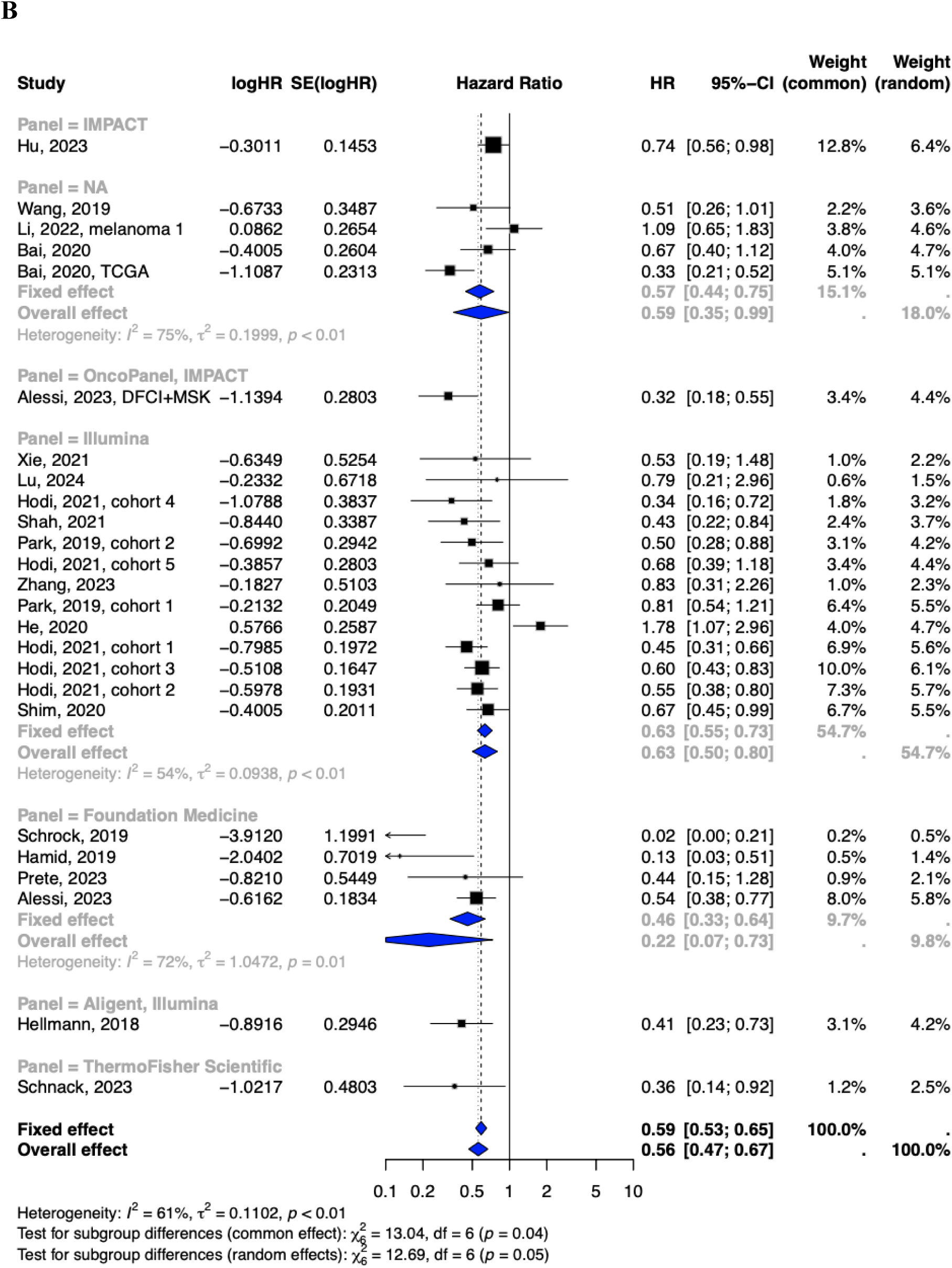
Subgroup analyses of OS and PFS by sequencing panels and approaches. (**A, B**) OS and PFS analyses stratified by sequencing panels and assays show moderate to high heterogeneity across commercial platforms. Despite these discrepancies, high TMB consistently correlated with better OS and PFS outcomes across all sequencing protocols, reaffirming its value as a predictive biomarker. These results underscore the importance of standardising sequencing methods and adopting high-coverage approaches, such as NGS, to improve reproducibility and reliability in TMB assessment.

### Publication bias and sensitivity analysis

Publication bias was evaluated using funnel plots (**Supplementary Figure 1A, 1B**), which displayed symmetry, suggesting that the included studies are unlikely to be influenced by publication bias. Egger’s test further supported these findings, revealing minimal bias for OS studies (*p* = 0.0086) and no significant bias for PFS studies (*p* = 0.0735). The trim-and-fill method applied to OS studies confirmed these results, showing no evidence of publication bias (*p* = 0.8195) (**Supplementary Figure 2**).

## Discussion

This meta-analysis of 5,278 patients from 28 studies offers critical insights into the clinical utility of TMB as a predictive biomarker for survival outcomes and treatment stratification. By aggregating survival data from high and low TMB cohorts, our findings demonstrate that high TMB is consistently associated with significant improvements in OS and PFS. These results underscore the central role of TMB in predicting treatment responses despite inherent heterogeneity across studies.

Subgroup analyses revealed high TMB’s strong association with better OS across cancer types, particularly in NSCLC and GI cancers, and in advanced or metastatic stages, consistent with prior findings [41, 42]. This correlation reflects the immunogenic potential of neoantigens produced during tumour progression. As somatic mutations accumulate, they generate a repertoire of neoantigens that facilitate tumour-specific immune responses, enhancing the efficacy of ICI therapies and ultimately improving the OS outcomes across cancer types in ICI-treated cohorts [43]. Importantly, we found that patients receiving anti-PD-L1/PD-1 therapies experienced the most pronounced survival benefits for both OS and PFS, suggesting the pivotal role of these inhibitors in high TMB tumours. Combined checkpoint blockade therapies targeting PD-L1/PD-1 and CTLA-4 yielded even stronger outcomes, likely due to their ability to overcome diverse immune escape mechanisms. While these findings highlight TMB’s predictive utility in immunotherapy, the moderate-to-high heterogeneity observed in anti-CTLA-4 cohorts underscores the need for more focused studies with larger sample sizes to validate these results.

The association of high TMB with improved survival in chemotherapy-treated cohorts showed greater variability, reflecting the complex interplay of genomic instability, treatment resistance, and tumour biology. High TMB may amplify chemotherapy sensitivity by increasing genomic instability, leading to a longer, durable period. On the other hand, it can also facilitate chemoresistance through additional mutational burdens that impair drug efficacy [44, 45]. Our meta-analysis revealed a better prognosis associated with the predictive value of high TMB, reflected in improved OS and PFS. Our study’s limited number of chemotherapy-treated cohorts restricted a more nuanced analysis of chemotherapy subtypes, indicating the need for further investigation into TMB’s role in predicting responses to specific chemotherapeutic agents.

The predictive value of TMB also varied by tumour stage, influenced by factors such as immune cell infiltration and neoantigen quality. Early-stage tumours with high TMB exhibited more immunogenic neoantigens, enhancing immune recognition, infiltration, and therapeutic responses [8, 46]. This aligns with the stronger association between high TMB and prolonged PFS in primary tumours. In contrast, advanced and recurrent tumours are characterised by immune evasive strategies, including immune editing and the production of less immunogenic neoantigens due to subclonal diversity [8, 47]. These dynamics may explain the diminished PFS benefit of high TMB in advanced stages. Similarly, despite being primary tumours, gliomas and penile SCC behave aggressively with profound immune suppression and significant intratumoural heterogeneity. These factors lead to worse outcomes even in the presence of high TMB. Conversely, high TMB is associated with better OS in relatively immunocompetent cancer types, likely due to the favourable immune infiltration and desirable neoantigen quality that effectively stimulate anti-tumour responses. These findings highlight the importance of understanding tumour stage and microenvironmental factors when interpreting TMB as a biomarker. The variability in TMB quantification across different sequencing platforms underscores a significant challenge in standardising TMB as a predictive biomarker. Differences in gene coverage, sequencing depth, and mutation detection sensitivity among commercial platforms (e.g., IMPACT, Foundation Medicine, MSKCC, and Illumina) contribute to heterogeneity in survival analyses, particularly in OS and PFS outcomes. Despite these discrepancies, the consistent association between high TMB and improved survival across all platforms reinforces its prognostic value. These findings highlight the need for standardised sequencing protocols and the adoption of high-coverage approaches to enhance reproducibility and ensure more reliable clinical decision-making based on TMB assessment.

Despite the comprehensive scope of this analysis, certain limitations warrant consideration. The small sample sizes in certain treatment groups, particularly for anti-CTLA-4 therapies and chemotherapy, reduced statistical power and generalisability. Additionally, the variability introduced by diverse sequencing platforms, with differences in coverage, depth, and accuracy, could affect the robustness of TMB as a biomarker. Finally, the assumption that high TMB universally enhances immunogenicity may not hold, as high TMB tumours can produce poorly immunogenic neoantigens, contributing to worse outcomes in certain cancer types with unique tumour microenvironments. Furthermore, specific cancer types, distinct molecular subtypes and varying TMB levels and therapeutic responses highlight the need for cancer-type-specific analysis. Such analyses will become increasingly feasible as larger cohorts become available in the future.

Addressing these limitations requires future research to prioritise the standardisation of TMB measurement protocols and the establishment of clinically relevant thresholds. Large-scale, randomised trials are essential to validate TMB’s predictive value across diverse treatment modalities. Integrating complementary biomarkers, such as neoantigen quality, immune cell infiltration, and immune checkpoint expression, could further enhance the accuracy of TMB in predicting treatment responses. Multi-omic approaches that combine TMB assessment with transcriptomics, proteomics and immune profiling will refine our understanding of TMB’s role in guiding personalised cancer therapies. This comprehensive framework will strengthen the use of TMB as a robust biomarker, ensuring its consistency and reliability in clinical decision-making.

## Author Contributions

Study concept and design: AM (Aijia Meng), AY, HWS, ST, HN, JY, AM (Ashish Mehta), JP, AZ. Data curation: AM (Aijia Meng), AZ. Data analysis and interpretation: AM (Aijia Meng), AY, HWS, ST, HN, JY, AM (Ashish Mehta), AZ. Writing-drafting original manuscript: AM (Aijia Meng), AZ. Writing-reviewing and editing: AM (Aijia Meng), AY, HWS, ST, HN, JY, AM (Ashish Mehta), JP, AZ. Study supervision: JP, AZ. All authors critically review the final manuscript for intellectual content.

## Data Availability

The data included in this meta-analysis are available in the article and online supplementary materials.

## Data Availability

All data produced in the present work are contained in the supplementary materials.

