## Supplementary Figures for "A Systematic Review and Meta-Analysis of the Impact of Tumour Mutation Burden on Survival Outcomes in Solid Tumours"

**Literature** **search** **strategies**
We systematically searched the relevant literature in PubMed, Scopus, ScienceDirect and Cochrane databases from 01/01/2010 to 14/08/2024, with subject headings and free worlds search method. The following keywords were used: TMB, tumour mutation burden, tumor mutation burden, tumour mutational burden, tumor mutational burden, overall survival, and progression-free survival.

**A**	**B**

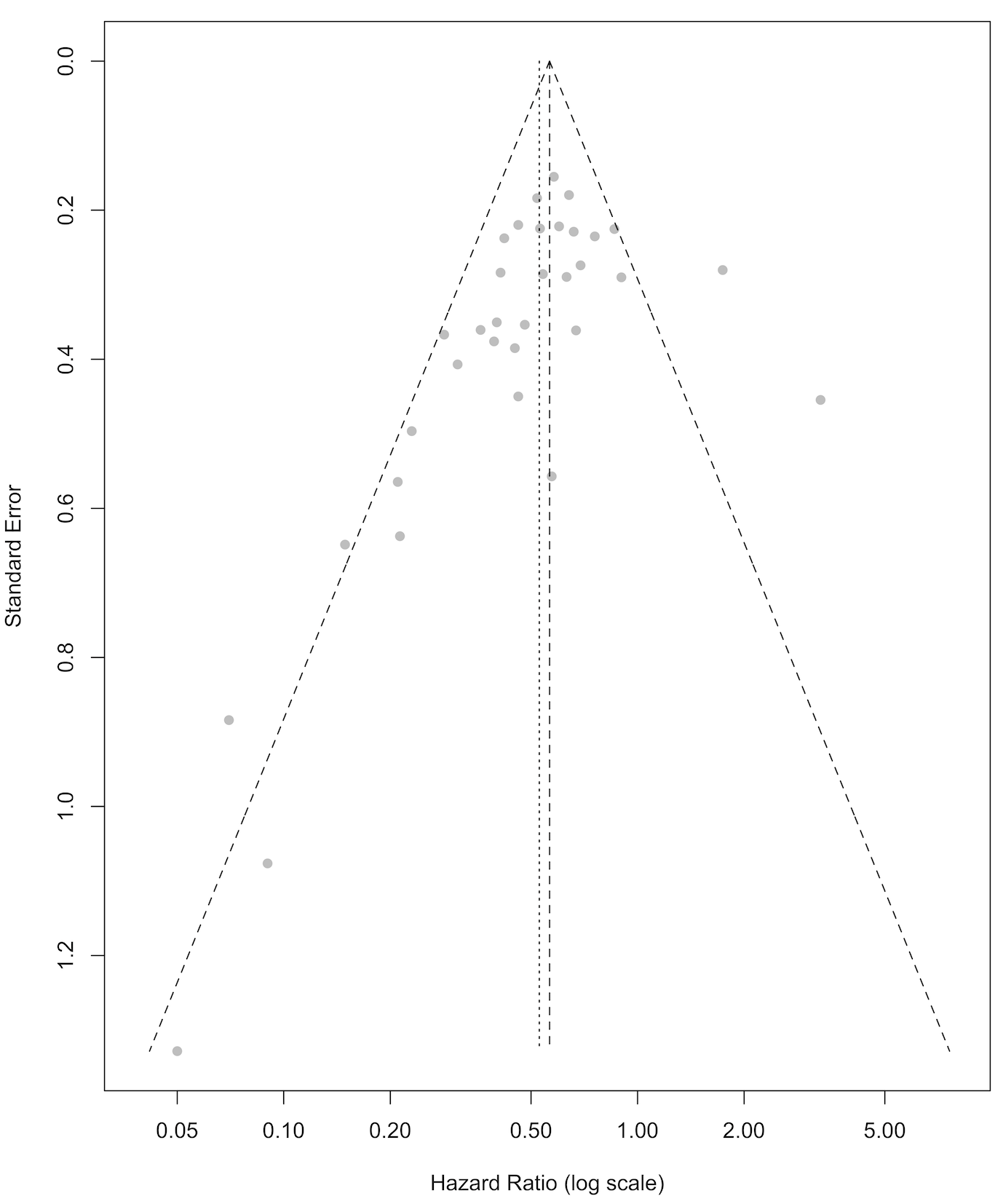

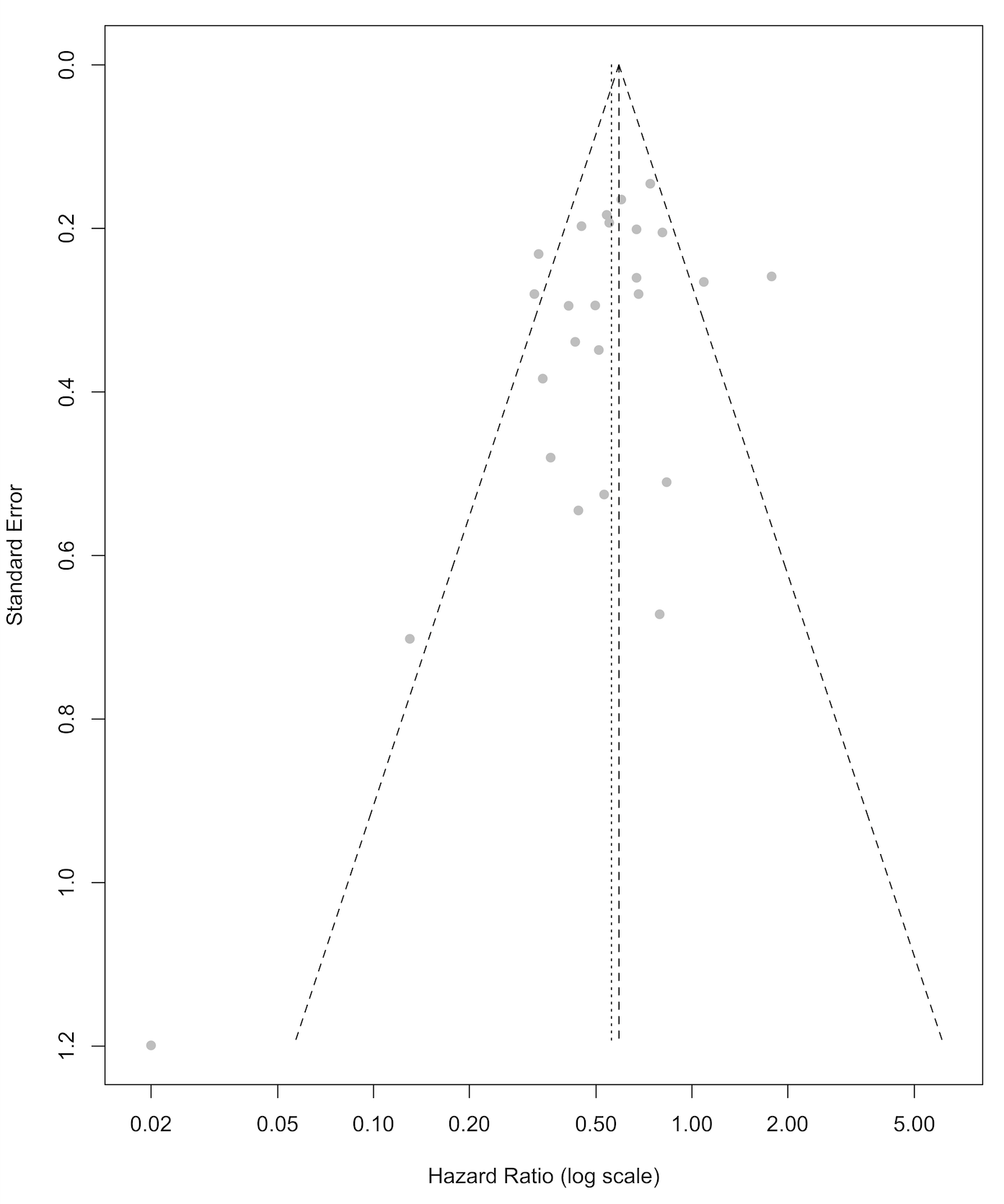

**Supplementary Figure 1. Funnel plots with pseudo 95% CI for a) OS and b) PFS.** The plots showed horizontal scatters, consistent with the meta-analyses that pooled results from selected studies.

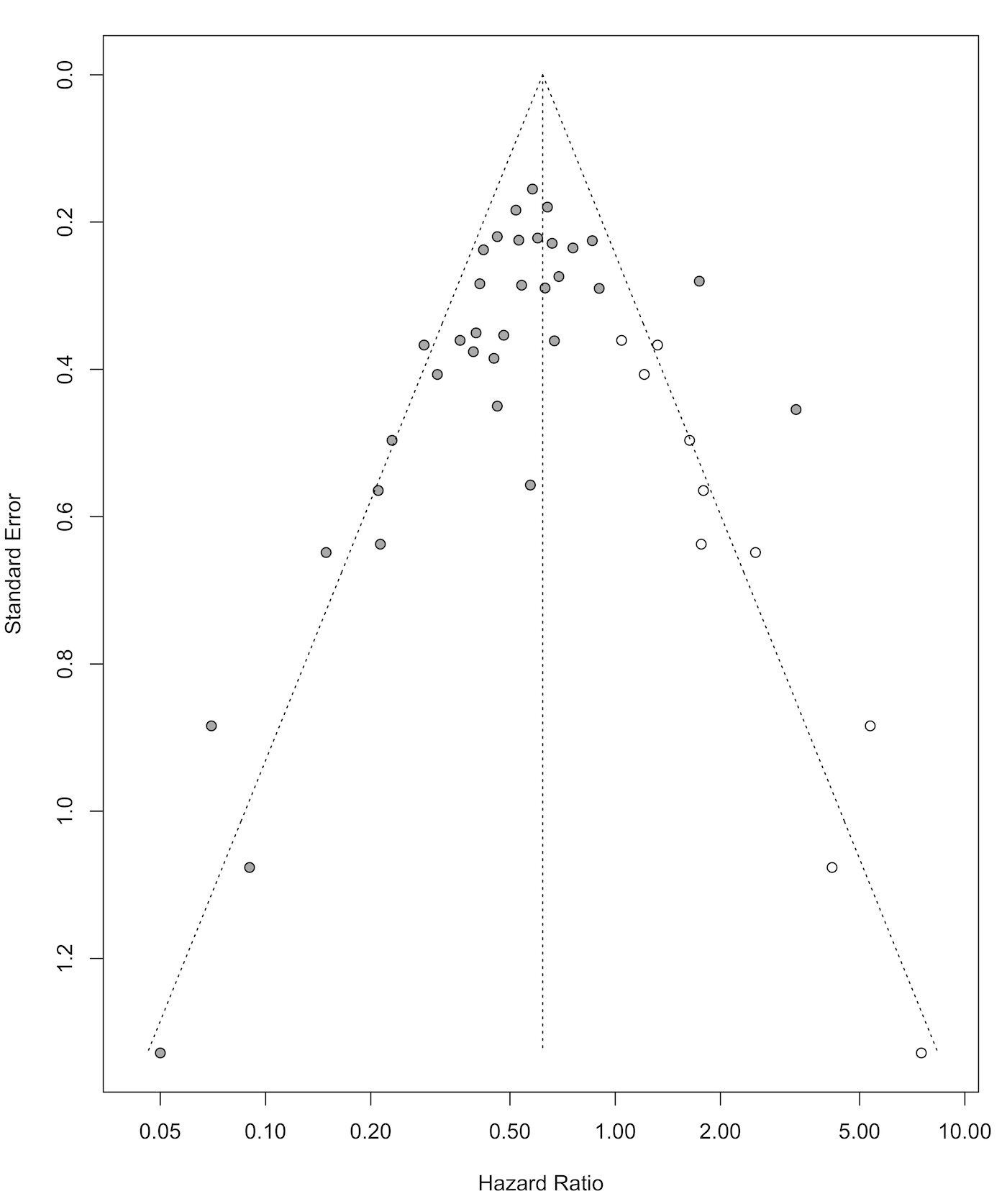

**Supplementary Figure 2. Funnel plot for OS studies after trim-and-fill analysis.** Grey dots represent the studies originally included in this meta-analysis. White dots indicate the imputed studies that were corrected from the trim-and-fill method. Overall, the funnel plot is symmetric, suggesting no evidence of publication bias.
